# Sleeping posture, behaviour, and environment in late pregnancy: A comprehensive analysis of a video-based, multi-night, in-home, level 3 sleep apnea study of pregnant participants and their bed partners

**DOI:** 10.64898/2026.02.03.26345492

**Authors:** Allan J Kember, Leah Ritchie, Hafsa Zia, Praniya Elangainesan, Noa Gilad, Jane Warland, Babak Taati, Elham Dolatabadi, Sebastian R Hobson

## Abstract

To characterize sleeping posture, behaviour, and environment in healthy pregnant participants and their bed partners across multiple nights in the home setting during late pregnancy, we completed a manual review of overnight video recordings from a four-night, in-home, level 3 sleep apnea study. Sleeping postures were scored according to a thirteen-posture classification system to determine the cumulative time per night spent in each sleeping posture. Additional aspects of sleeping posture, behaviour, and environment were also assessed. Forty-one pregnant participants and 36 bed partners completed the study, contributing 168 nights of video. Significant differences were found between the pregnant participants and bed partners in cumulative time spent in each posture as well as frequency and duration of episodes spent in each posture. Pregnancy status, side of the bed, and presence of a pregnancy pillow, bed partner, shared bed sheets, and pets in the sleeping space had various effects on the time spent in each posture. Pregnant participants spent more time in transition postures (going-to-sleep, waking-to-void, returning-to-bed, and waking-in-the-morning) than bed partners. There was a moderately positive correlation in posture changes between pregnant participants and their bed partners. Pets significantly increased the number of posture changes per night for both groups. Pregnant participants had more absences and time absent from bed. Sleep in late pregnancy is characterized by an increased frequency and duration of episodes spent in a restricted number of sleeping postures and is impacted by the sleep environment. Modifying the sleeping environment may improve comfort, minimize disturbances, and benefit sleep.

**Statement of Significance:** Subjectively-recalled supine going-to-sleep posture in late pregnancy is associated with stillbirth and fetal growth restriction. Sleeping posture, however, is dynamic, and few studies provide comprehensive analyses of sleeping posture in pregnancy using objective measurements. This novel study used a gold-standard objective measure of sleeping posture, was conducted across multiple nights in the participant’s own homes, and accounts for usual sleeping behaviours and environment by including the participant’s bed partner. A critical remaining knowledge gap is whether an individual’s nightly sleeping posture varies significantly across the third trimester. Future work should use nightly, continuous, and objective methods to measure sleeping posture across the entire third trimester to bridge this gap and investigate the relationship between sleeping posture and pregnancy outcomes.

## Introduction

Sleep is an essential component of health and wellbeing; it consumes approximately one-third of human life, yet poor sleep can critically compromise the other two-thirds.^1,2^ Accumulating evidence highlights that sleep disorders are common in pregnancy, and remain largely underdiagnosed, undertreated, and with direct links to pregnancy complications.^3–8^

Following a landmark study in 2011,^9^ maternal supine going-to-sleep posture in late pregnancy has gained attention as a possible contributor to adverse pregnancy outcomes including late stillbirth and giving birth to a growth-restricted infant.^10–16^ When the supine posture is assumed in late pregnancy, the gravid uterus compresses the retroperitoneal vasculature,^17–21^ which can result in reduced venous return, reduced preload, reduced cardiac output, and reduced blood flow in the common and internal iliac arteries and to the maternal side of the placenta.^18–23^ These alterations in maternal hemodynamics are reflected in placental and fetal hemodynamics,^19,24–29^ oxygen transfer to the fetus,^19,30,31^ and fetal behavioural state,^22^ and are proposed as one possible biologic mechanism linking maternal supine posture to adverse pregnancy outcomes. Furthermore, it is well-established that the supine posture exacerbates snoring and obstructive sleep apnea (OSA),^32^ and this has also been demonstrated in pregnancy with the prevalence of posture-dependent OSA recently reported as 43% in the third trimester.^33–36^ For further details, readers are directed to three recent comprehensive reviews of the impact of maternal posture on maternal physiology,^37^ fetal physiology,^38^ and maternal-fetal pathophysiology.^39^

The aforementioned studies demonstrating an association between maternal supine going-to-sleep posture in late pregnancy and adverse pregnancy outcomes were retrospective and going-to-sleep and waking postures were subjectively reported by the participants rather than objectively measured (e.g., video, accelerometry). As such, these studies are limited by potential for recall bias, established inaccuracies inherent in self-reported sleep behaviour when compared to objective measurements,^40–43^ and by the fact that sleeping position is not static.^43,44^ According to a recent review,^38^ there have been nine studies in participants in the third trimester in which sleeping posture was objectively verified.^34,35,45–51^ Of these, the largest and most comprehensive was by Wilson et al.^35^ who confirmed that, for most pregnancies, the going-to-sleep and waking postures are not representative of sleeping posture occurring throughout the night. Their study yields unprecedented clinical insight into sleeping posture in pregnancy and was the first to demonstrate a negative correlation between objectively-determined proportion of time spent supine overnight and birth weight.^35^

Wilson et al.’s study, however, was conducted over a single night,^35^ which does not account for night-to-night sleep variability. It was conducted in various locations (hospital, sleep laboratory, or home) depending on participant circumstances and preferences. It classified sleeping posture according to a five-posture system (left, right, prone, supine, upright), which does not account for tilted or twisted postures impacting uteroplacental hemodynamics. It used video verification of posture only in their attended sleep laboratory studies (21.4% of their studies) and did not account for the presence of a bed partner and other sleep environment variables (e.g., bed sheets, pillows, pets, children). As such, there remains a need for multi-night, in-home, sleep studies that employ objective sleeping posture determination (preferably video-based) accounting for unique aspects of pregnancy physiology as well as the sleep environment.

In the present study, we present a sub-analysis of the SLeeP AID4 (Sleep in Late Pregnancy: Artificial Intelligence Development for the Detection of Disturbances and Disorders) Study,^52^ which was a Canada-wide, video-based, four-night, in-home, level 3 sleep apnea study of pregnant participants and their bed partners in the third trimester. The aim of the present study was to jointly analyze continuous video of healthy pregnant people and their bed partners across four nights in the home setting to capture sleeping posture dynamics, transitional behaviours, and environmental interactions that are not observable in laboratory settings or short-duration recordings.

## Methods

### Design

Data analyzed were derived from a prospective, observational, video-based, four-night, in-home, level 3 sleep apnea study in pregnant people and their bed partners (ClinicalTrials.gov Identifier: NCT05376475).

### Participants

Participants were recruited Canada-wide using various social media platforms. All data were collected in the participant’s home. Inclusion criteria for participants included having an American Society of Anesthesiologists Physical Status (ASA PS) class II or lower,^53^ having a low-risk singleton pregnancy, being in the third trimester (28 weeks and 0 days through 40 weeks and 6 days, inclusive), being aged 18 to 50 years, having a 2.4 GHz Wi-Fi network in the home, and sleeping in a bed at night. Inclusion criteria for bed partners included having an ASA PS class was II or lower and sleeping in the same bed as the pregnant participant. Participants and bed partners were excluded if they were non-English speaking, reading, or writing. Bed partners were given the option to decline participation in the study. If they chose this option, they slept in a different room on the study nights.

### Interventions

Voluntary, written, informed consent was obtained from each participant (and bed partner, if participating). At entry to the study, we interviewed participants (and bed partners, if participating) to collect demographic and anthropometric information. We mailed the study equipment to each participant and met with them (and their bed partner, if participating) virtually to provide orientation to the study protocol and assistance in setting up the equipment. Participants attached an app-enabled home-surveillance video camera (Wyze Cam V2 by Wyze Labs, Inc., Seattle, USA) to the wall, centred at the head of their bed, and located 1.6 to 1.7 metres above the sleeping surface so that it clearly captured all pertinent sleep posture data. We provided a study iPhone (iPhone 7 by Apple Inc., Cupertino, USA) with the Wyze App installed, which the participant used to record continuous overnight video recordings directly to its hard drive. Participants started the video recording immediately prior to going to sleep each night and stopped the recording immediately upon waking each morning. As such, participants had full control over their video recordings, including the ability to permanently delete them. Also immediately prior to going to sleep, each participant (and their bed partner, if participating) donned two home sleep apnea sensors (NightOwl by Ectosense NV, Leuven, Belgium). One sensor was taped to the index finger and recorded peripheral oxygen saturation, pulse rate, photoplethysmography, and motion via accelerometry. The other sensor was taped just below the xiphoid process on the abdomen and recorded body posture via accelerometry. Each participant (and their bed partner, if participating) completed this data collection process for four nights. Participants (and their bed partner, if participating) were instructed to sleep according to their usual patterns and behaviours including sleeping attire, lighting, and bed sheet and pillow use. Upon completion of the study, we met virtually with each participant (and their bed partner, if participating) for an exit interview regarding their experience in the study. Following birth, we followed up with each participant to collect birth outcomes.

### Outcomes

The primary outcome of this study was the video-based cumulative length of time per night and cumulative percentage of the night spent in each sleeping posture for the pregnant participant and bed partner, if participating. Sleeping posture for the participant and bed partner was defined according to thirteen previously described sleeping postures (prone, left recovery, left lateral, left tilt, supine, supine thorax with left pelvic tilt, supine thorax with right pelvic tilt, supine pelvis with left thorax tilt, supine pelvis with right thorax tilt, right tilt, right lateral, right recovery, and sitting; see **Figure 1**).^52,54^ Given that the position of the pelvis impacts maternal, placental, and fetal hemodynamics and the position of the thorax impacts OSA physiology,^37,38^ the thirteen-posture classification system was previously defined in order to account for the position of the pelvis and thorax independently. Each participant’s and bed partner’s overnight video was manually reviewed and sleeping posture was scored according to the classification system for each 30-second epoch for the entire duration of the video. Video review and scoring was completed by two trained research personnel (AJK and LR). When two or more postures occurred within one epoch, the epoch’s posture was scored as the posture that occupied the majority of the epoch.

**Figure 1.**
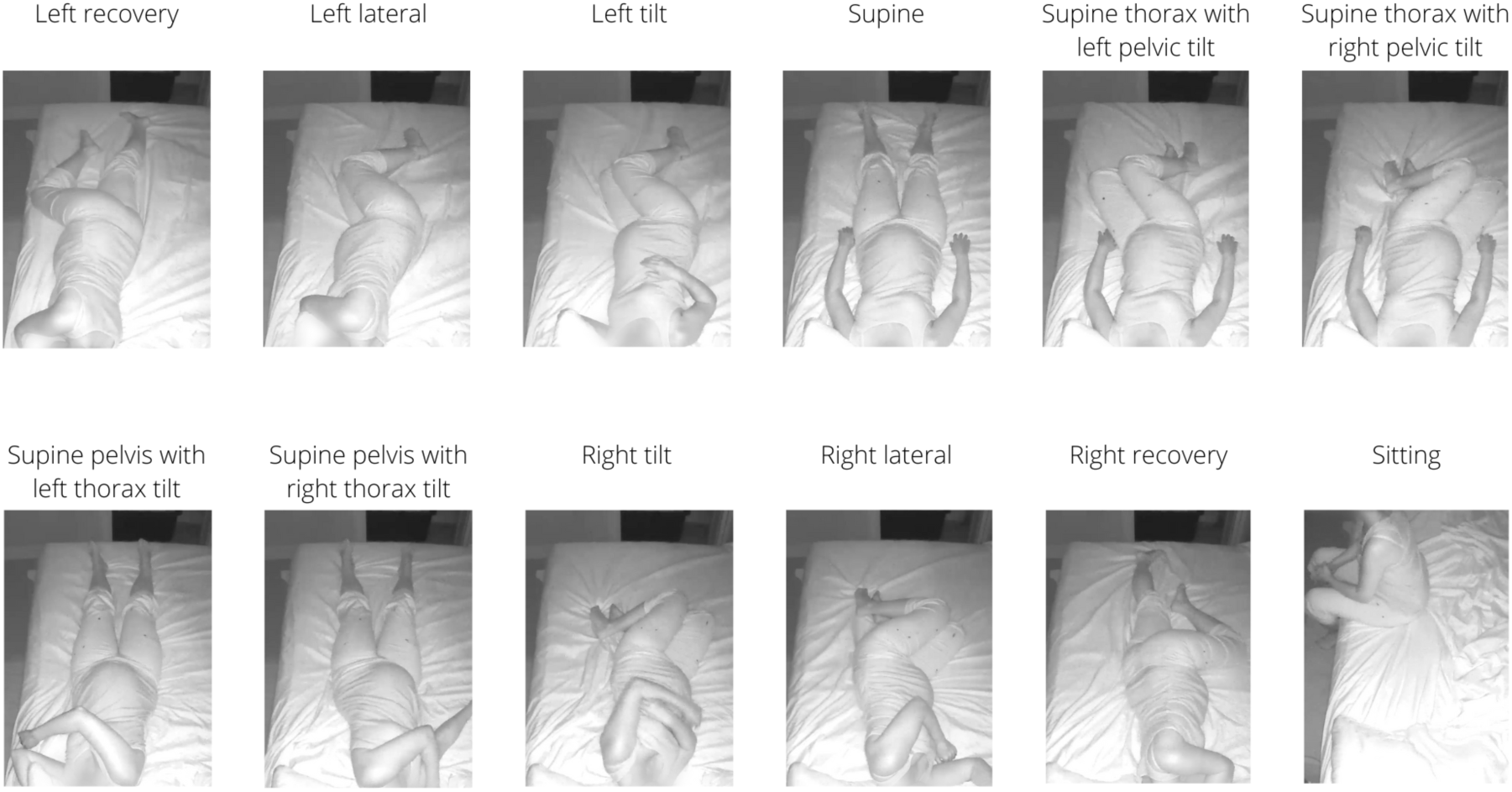
Thirteen-posture sleeping posture classification system demonstrating examples of twelve previously defined sleeping postures.^52,54^ Note that the prone posture is not shown. This figure was adapted with permission from the SLeeP AIDePt study.^54^ Permission was granted by the author (AJK).

Secondary outcomes for the pregnant participant and bed partner included additional aspects of sleep posture, sleep behaviour, and sleep environment variables. Additional aspects of sleep posture included: the number of episodes spent in each posture overnight; the average and maximum duration of episodes spent in each posture overnight; the immediate and cumulative nightly amount of time spent in transition postures; the total number of posture change events per night; the total number of posture changes per person per night; posture change interactions between the participant and bed partner; and the total and hour-by-hour posture change index. Sleep behaviour included the total number of absences per night and the total time spent absent from bed per night. We defined an absence as an event in which the person exited the bed for the majority of an epoch (at least 15 seconds) and then subsequently returned to the bed. In the event that a person came to bed late (e.g., stayed up later than their co-sleeper) or left the bed early (e.g., got up earlier than their co-sleeper to go to work), this was not counted as an absence. During scoring of each participant’s overnight video, sleep environment variables, including the side of the bed the participant slept on, whether bedsheets were shared or separate, and the presence of a bed partner, pregnancy pillows, pets, and children were noted.

The NightOwl-based sleeping posture data, which follows a five-posture classification system (i.e., left, right, prone, supine, upright) was not used in this study except in rare circumstances where video-based posture classification was difficult (e.g., obscuration of the participant’s or bed partner’s posture by large pillows or bulky duvets), and it was only reviewed to assist in clarification of the video-based posture. The NightOwl-based sleeping posture data is presented and analyzed in the **Supplementary File**. Furthermore, note that the NightOwl sleep physiology data (peripheral oxygen saturation, pulse rate, photoplethysmography), exit interview data, and birth outcome data is beyond the scope of the present report and will be published separately.

### Sample Size

In logistic regression analyses, guidelines developed by Peduzzi et al.^55^ and Concato et al.^56^ indicate that approximately ten participants are required per each independent variable of interest. In the SLeeP AID4 study, six variables of interest relating to the pregnant participant were selected (one of which was sleeping posture), so a target sample size of n = 60 couples (60 pregnant participants and 60 bed partners) was selected.

### Statistical Methods

Statistical analyses were conducted using JASP (Version 0.95.1) and R (Version 4.4.2). Normality of continuous variables was assessed using the Shapiro-Wilk test at a 0.05 significance level. Normally distributed continuous variables are presented as mean ± standard deviation, whereas non-normally distributed continuous variables are presented as median (interquartile range). A Student T-test was used for difference testing of continuous variables if the variable was normally distributed in both groups, whereas a Mann-Whitney test was used if the variable was non-normally distributed in one or both groups. A statistically significant difference was indicated by a two-sided p-value of less than 0.05. Multiple linear regression was used to explore the impact of sleep environment variables on sleeping posture and sleep behaviours for the pregnant participants and their bed partners. Pearson correlation coefficient was calculated to evaluate the synchrony of postural changes within couples, using night-wise posture change data.

### Ethical Approval

This study was conducted in accordance with The Tri-Council Policy Statement: Ethical Conduct for Research Involving Humans and was approved by the University of Toronto Health Sciences Research Ethics Board (Protocol No. 41612).

## Results

A total of 128 people expressed interest in participating in the SLeeP AID4 Study from July 2022 through April 2024. Of these, forty-four (33%) participants were screened, met the eligibility criteria, were recruited, and gave voluntary, written, informed consent. Of these 44 participants, one did not have a bed partner, five bed partners declined participation, and the remaining 38 bed partners were screened, met eligibility criteria, were recruited, and gave voluntary, written, informed consent. Three participants and two bed partners dropped out before commencing the study. A total of 41 pregnant participants and 36 bed partners successfully completed the study, which means that we were underpowered for some analyses as discussed in the Limitations section.

### Demographics & Anthropometrics

Demographic and anthropometric characteristics of the study sample are published separately.^57^

### Sleep Environment Variables

During study participation, thirty-six (87.8%) of pregnant participants slept with a bed partner, 22 (53.7%) slept on the left side of the bed (as viewed from the perspective of a person sitting in the bed), eleven (26.8%) had pets present on the bed, six (14.6%) had a child or children enter their bedroom at one or more points in the night, and eighteen (43.9%) slept using a pregnancy/body pillow. Twenty eight couples (of 36 total; 77.8%) slept under the same set of bedsheets (i.e., they shared the set of bedsheets and they did not use separate sets of bedsheets). Thirty-seven pregnant participants (90.2%) completed the study in the antepartum period, while three (7.3%) gave birth after recruitment but prior to commencing the first night of the study and, therefore, completed the study in the postpartum period. One participant (2.4%) gave birth after the first night of the study and subsequently completed the second through fourth night of the study in the postpartum period.

### Sleeping Posture Characterization

In the following sections, we present the results of a manual review and posture scoring of 168 nights (71,010 minutes; 142,020 30-second epochs) of overnight video.

#### Cumulative Time Analysis

The cumulative number of minutes per night and cumulative percentage of the night spent in each posture by the pregnant participants and bed partners based on analysis of the overnight video recordings is shown in **Table 1**. Difference testing, the 95% confidence interval of the difference, and the p-value is also shown. Participants (and their bed partner, if applicable) who completed the sleep studies postpartum were excluded from this analysis and are presented separately in the **Supplementary File**.

**Table 1.**
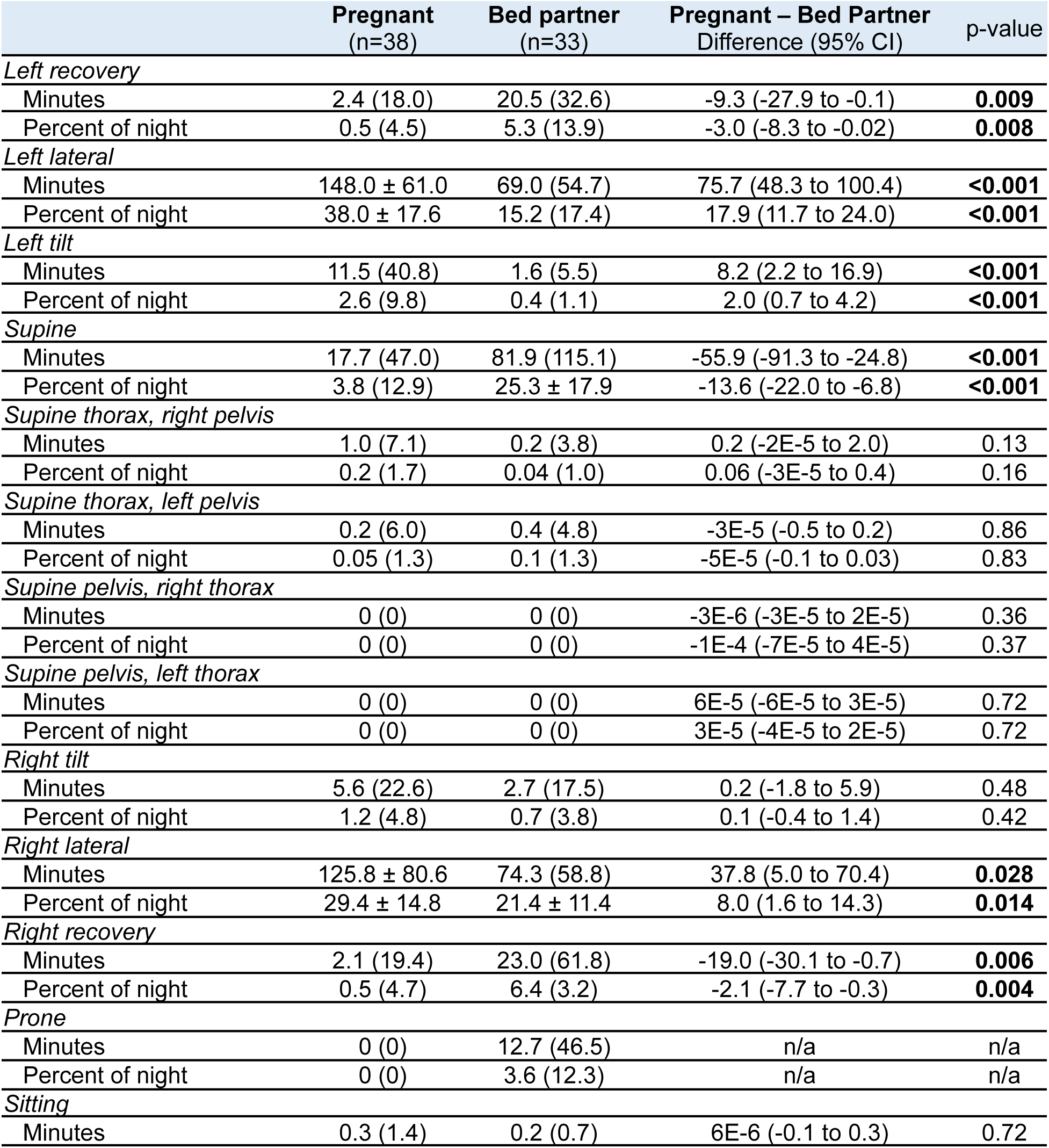
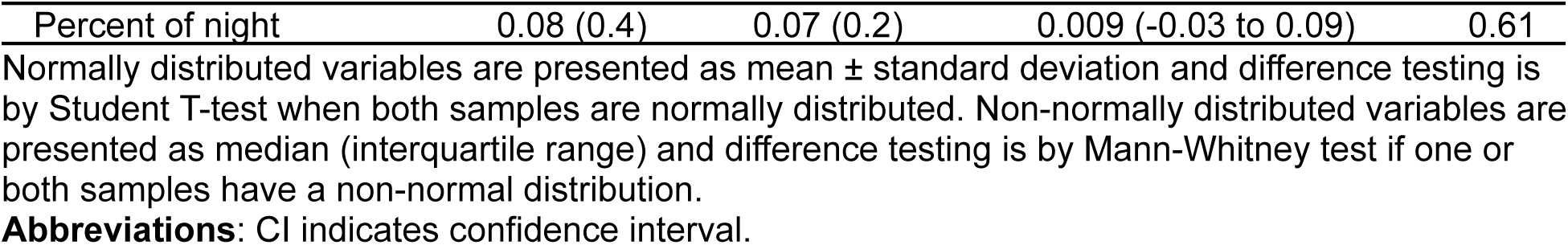
Cumulative number of minutes per night and cumulative percentage of the night spent in each posture for the pregnant participants and bed partners based on video analysis.

|  | Pregnant<br>(n=38) | Bed partner<br>(n=33) | Pregnant – Bed Partner<br>Difference (95% CI) | p-value |
| --- | --- | --- | --- | --- |
| *Left recovery* |  |  |  |  |
| Minutes | 2.4 (18.0) | 20.5 (32.6) | -9.3 (-27.9 to -0.1) | **0.009** |
| Percent of night | 0.5 (4.5) | 5.3 (13.9) | -3.0 (-8.3 to -0.02) | **0.008** |
| *Left lateral* |  |  |  |  |
| Minutes | 148.0 ± 61.0 | 69.0 (54.7) | 75.7 (48.3 to 100.4) | **<0.001** |
| Percent of night | 38.0 ± 17.6 | 15.2 (17.4) | 17.9 (11.7 to 24.0) | **<0.001** |
| *Left tilt* |  |  |  |  |
| Minutes | 11.5 (40.8) | 1.6 (5.5) | 8.2 (2.2 to 16.9) | **<0.001** |
| Percent of night | 2.6 (9.8) | 0.4 (1.1) | 2.0 (0.7 to 4.2) | **<0.001** |
| *Supine* |  |  |  |  |
| Minutes | 17.7 (47.0) | 81.9 (115.1) | -55.9 (-91.3 to -24.8) | **<0.001** |
| Percent of night | 3.8 (12.9) | 25.3 ± 17.9 | -13.6 (-22.0 to -6.8) | **<0.001** |
| *Supine thorax, right pelvis* |  |  |  |  |
| Minutes | 1.0 (7.1) | 0.2 (3.8) | 0.2 (-2E-5 to 2.0) | 0.13 |
| Percent of night | 0.2 (1.7) | 0.04 (1.0) | 0.06 (-3E-5 to 0.4) | 0.16 |
| *Supine thorax, left pelvis* |  |  |  |  |
| Minutes | 0.2 (6.0) | 0.4 (4.8) | -3E-5 (-0.5 to 0.2) | 0.86 |
| Percent of night | 0.05 (1.3) | 0.1 (1.3) | -5E-5 (-0.1 to 0.03) | 0.83 |
| *Supine pelvis, right thorax* |  |  |  |  |
| Minutes | 0 (0) | 0 (0) | -3E-6 (-3E-5 to 2E-5) | 0.36 |
| Percent of night | 0 (0) | 0 (0) | -1E-4 (-7E-5 to 4E-5) | 0.37 |
| *Supine pelvis, left thorax* |  |  |  |  |
| Minutes | 0 (0) | 0 (0) | 6E-5 (-6E-5 to 3E-5) | 0.72 |
| Percent of night | 0 (0) | 0 (0) | 3E-5 (-4E-5 to 2E-5) | 0.72 |
| *Right tilt* |  |  |  |  |
| Minutes | 5.6 (22.6) | 2.7 (17.5) | 0.2 (-1.8 to 5.9) | 0.48 |
| Percent of night | 1.2 (4.8) | 0.7 (3.8) | 0.1 (-0.4 to 1.4) | 0.42 |
| *Right lateral* |  |  |  |  |
| Minutes | 125.8 ± 80.6 | 74.3 (58.8) | 37.8 (5.0 to 70.4) | **0.028** |
| Percent of night | 29.4 ± 14.8 | 21.4 ± 11.4 | 8.0 (1.6 to 14.3) | **0.014** |
| *Right recovery* |  |  |  |  |
| Minutes | 2.1 (19.4) | 23.0 (61.8) | -19.0 (-30.1 to -0.7) | **0.006** |
| Percent of night | 0.5 (4.7) | 6.4 (3.2) | -2.1 (-7.7 to -0.3) | **0.004** |
| *Prone* |  |  |  |  |
| Minutes | 0 (0) | 12.7 (46.5) | n/a | n/a |
| Percent of night | 0 (0) | 3.6 (12.3) | n/a | n/a |
| *Sitting* |  |  |  |  |
| Minutes | 0.3 (1.4) | 0.2 (0.7) | 6E-6 (-0.1 to 0.3) | 0.72 |
| Percent of night | 0.08 (0.4) | 0.07 (0.2) | 0.009 (-0.03 to 0.09) | 0.61 |

Normally distributed variables are presented as mean ± standard deviation and difference testing is by Student T-test when both samples are normally distributed. Non-normally distributed variables are presented as median (interquartile range) and difference testing is by Mann-Whitney test if one or both samples have a non-normal distribution.
**Abbreviations:** CI indicates confidence interval.

#### Episodic Time Analysis

For each posture, the average number of episodes spent in that posture per night, the average length of these episodes, and the average maximum length of these episodes, is given in **Table 2** for the pregnant participants and bed partners based on analysis of the overnight videos. Participants (and their bed partner, if applicable) who completed the sleep studies postpartum were excluded from this analysis and are presented separately in the **Supplementary File**.

**Table 2.** Average number of episodes in each posture per night, average length (in minutes) of episodes spent in each posture, and average maximum length (in minutes) of episodes spent in each posture for the pregnant participants and bed partners based on video analysis.

|  | Pregnant<br>(n=38) | Bed partner<br>(n=33) | Pregnant – Bed Partner<br>Difference (95% CI) | p-value |
| --- | --- | --- | --- | --- |
| *Left recovery episodes* | [17] | [6] |  |  |
| Avg. number per night | 1.0 (3.1) | 2.6 ± 1.8 | -0.5 (-1.8 to 0.33) | 0.29 |
| Avg. length (minutes) | 11.1 (10.5) | 14.9 ± 8.5 | -0.63 (-6.2 to 4.5) | 0.87 |
| Avg. max. length (minutes) | 29.5 (37.0) | 35.6 ± 21.3 | 1.0 (-14.5 to 15.0) | 0.87 |
| *Left lateral episodes* | [1] |  |  |  |
| Avg. number per night | 4.8 (2.7) | 3.7 (3.0) | 1.5 (0.50 to 2.5) | **0.006** |
| Avg. length (minutes) | 32.7 ± 15.2 | 21.4 ± 9.7 | 11.3 (5.3 to 17.3) | **< .001** |
| Avg. max. length (minutes) | 93.3 ± 35.8 | 53.5 (23.5) | 33.5 (19.5 to 51.5) | **< .001** |
| *Left tilt episodes* | [5] | [11] |  |  |
| Avg. number per night | 1.6 (3.3) | 0.62 (1.1) | 0.80 (0.25 to 2.0) | **0.003** |
| Avg. length (minutes) | 8.0 (7.0) | 6.9 (6.6) | 2.0 (-1.3 to 5.2) | 0.25 |
| Avg. max. length (minutes) | 24.5 (42.5) | 10.5 (14.8) | 12.5 (3.0 to 25.5) | **0.01** |
| *Supine episodes* | [3] |  |  |  |
| Avg. number per night | 2 (2.2) | 4.6 ± 2.4 | -1.9 (-0.75 to -2.9) | **< .001** |
| Avg. length (minutes) | 11.3 (17.4) | 20.5 (14.3) | -7.3 (-13.0 to -1.9) | **0.01** |
| Avg. max. length (minutes) | 36 (62.8) | 74.0 (74.0) | -35.0 (-54.5 to -9.5) | **0.006** |
| *STRP episodes* | [12] | [15] |  |  |
| Avg. number per night | 0.71 (1.2) | 0.37 (0.77) | 0.084 (-0.10 to 0.42) | 0.42 |
| Avg. length (minutes) | 6.1 (13.6) | 5.0 (8.4) | 1.5 (-1.8 to 5.5) | 0.40 |
| Avg. max. length (minutes) | 16.3 (18.0) | 8.5 (12.1) | 4.0 (-3.5 to 12.0) | 0.26 |
| *STLP episodes* | [15] | [14] |  |  |
| Avg. number per night | 1.0 (0.95) | 0.67 (0.36) | 0.19 (-0.25 to 0.64) | 0.40 |
| Avg. length (minutes) | 2.2 (7.0) | 4.7 (6.8) | -1.8 (-4.8 to 0.67) | 0.18 |
| Avg. max. length (minutes) | 5.5 (27.5) | 10 (25.5) | -3.5 (-10.0 to 3.5) | 0.27 |
| *SPRT episodes* | [33] | [26] |  |  |
| Avg. number per night | 0.33 (0.08) | 0.27 ± 0.12 | 0.083 (-0.083 to 1.9) | 0.19 |
| Avg. length (minutes) | 3.9 ± 4.6 | 5.7 ± 4.3 | -1.7 (-7.8 to 4.3) | 0.53 |
| Avg. max. length (minutes) | 8.5 ± 11.8 | 6.8 ± 5.1 | 1.7 (-12.7 to 16.1) | 0.77 |
| *SPLT episodes* | [31] | [28] |  |  |
| Avg. number per night | 0.29 (0.50) | 0.23 ± 0.05 | 0.083 (-7.5E-5 to 1.0) | 0.08 |
| Avg. length (minutes) | 2.3 (7.3) | 4.0 ± 2.2 | -1.0 (-4.5 to 17.5) | 0.46 |
| Avg. max. length (minutes) | 3 (15.8) | 3.5 (1.5) | -0.5 (-5.0 to 44) | 0.81 |
| *Right tilt episodes* | [7] | [11] |  |  |
| Avg. number per night | 1.3 (1.3) | 1.3 (1.5) | 0.060 (-0.50 to 0.65) | 0.77 |
| Avg. length (minutes) | 7.1 (13.6) | 7.6 (8.5) | -1.6 (-4.9 to 3.0) | 0.36 |
| Avg. max. length (minutes) | 22.0 (41.0) | 16.8 (31.1) | -2.5 (-12.5 to 12) | 0.632 |
| *Right lateral episodes* | [1] |  |  |  |
| Avg. number per night | 4.5 ± 2.1 | 3.9 ± 2.1 | 0.52 (-0.47 to 1.5) | 0.30 |
| Avg. length (minutes) | 31.4 (24.0) | 21.3 (10.1) | 7.6 (1.4 to 14.9) | **0.02** |
| Avg. max. length (minutes) | 79.0 (46.5) | 51.5 (23.0) | 23.5 (9 to 37.5) | **0.002** |
| *Right recovery episodes* | [15] | [3] |  |  |
| Avg. number per night | 1.3 (2.5) | 1.3 (1.3) | -0.071 (-1.2 to 0.42) | 0.71 |
| Avg. length (minutes) | 14.6 (14.1) | 14.8 (10.1) | 0.20 (-4.5 to 5.1) | 0.97 |
| Avg. max. length (minutes) | 37.2 ± 25.8 | 37.3 ± 26.5 | -0.078 (-14.6 to 14.5) | 0.99 |
| *Prone episodes* | [38] | [7] |  |  |
| Avg. number per night | 0 (0) | 1.0 (2.3) | n/a | n/a |
| Avg. length (minutes) | 0 (0) | 16.3 (14.1) | n/a | n/a |
| Avg. max. length (minutes) | 0 (0) | 37.3 ± 30.3 | n/a | n/a |
| *Sitting episodes* | [11] | [6] |  |  |
| Avg. number per night | 0.67 (1.1) | 0.50 (0.48) | 0.33 (2.3E-5 to 0.75) | **0.03** |
| Avg. length (minutes) | 0.80 (0.5) | 0.50 (0.90) | 7.5E-5 (-0.25 to 0.25) | 0.746 |
| Avg. max. length (minutes) | 1.0 (1.5) | 0.50 (2.5) | 3.4E-5 (-0.5 to 0.5) | 0.57 |

Normally distributed variables are presented as mean ± standard deviation and difference testing is by Student T-test when both samples are normally distributed. Non-normally distributed variables are presented as median (interquartile range) and difference testing is by Mann-Whitney test if one or both samples have a non-normal distribution. Square brackets with a number enclosed, [number], indicates the number of missing values (i.e., the number of participants that did not have any episodes in the posture).
**Abbreviations:** CI indicates confidence interval; Avg. indicates average; Max. indicates maximum; STRP indicates supine thorax, right pelvis; STLP indicates supine thorax, left pelvis; SPRT indicates supine pelvis, right thorax; SPLT indicates supine pelvis, left thorax.

#### Transition Postures Analysis

For each participant and bed partner, transition postures were analyzed to determine the length of time spent in the transition posture prior to changing to another posture (going-to-sleep posture, returning to bed posture) or since assuming that posture (waking-to-void posture, waking-in-the-morning posture) (see **Table 3**). Furthermore, for the going-to-sleep and waking-in-the-morning postures, the cumulative number of minutes per night and corresponding percentage of the night spent in each posture was determined. Participants (and their bed partner, if applicable) who completed the sleep studies postpartum were excluded from this analysis and are presented separately in the **Supplementary File**.

**Table 3.**
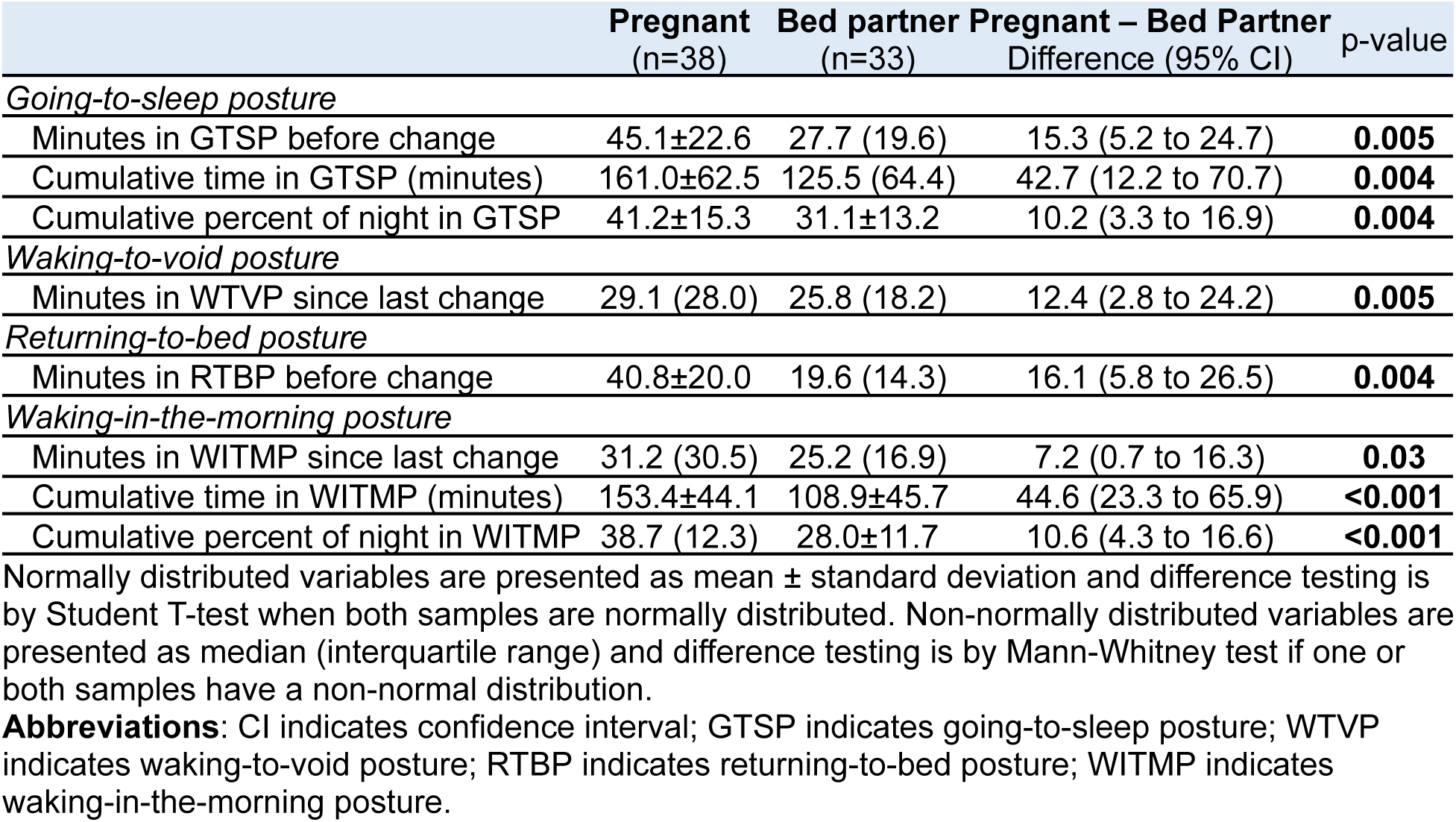
Immediate and cumulative nightly amount of time spent in transition postures for the pregnant participants and bed partners based on video analysis.

#### Sleeping Posture Changes Analysis

Overnight videos were analyzed to determine, for the 33 pregnant participant and bed partner couples, the average time in bed per night, the total number of posture change events (i.e., in which one or both co-sleepers changed posture) per night and the total number of posture changes per person per night (see **Table 4**). For each posture change event in the 33 couples, we indicated whether the co-sleeper did not change posture, changed posture within 30 seconds, 31-60 seconds, or 61-90 seconds. For the pregnant participant and bed partner, we provided the total nightly posture change index (PCI; the number of posture changes occurring per hour) and hour-by-hour PCI from the first hour through the last hour of the overnight video recordings to show how the frequency of posture changes varies throughout the night.

**Table 4.**
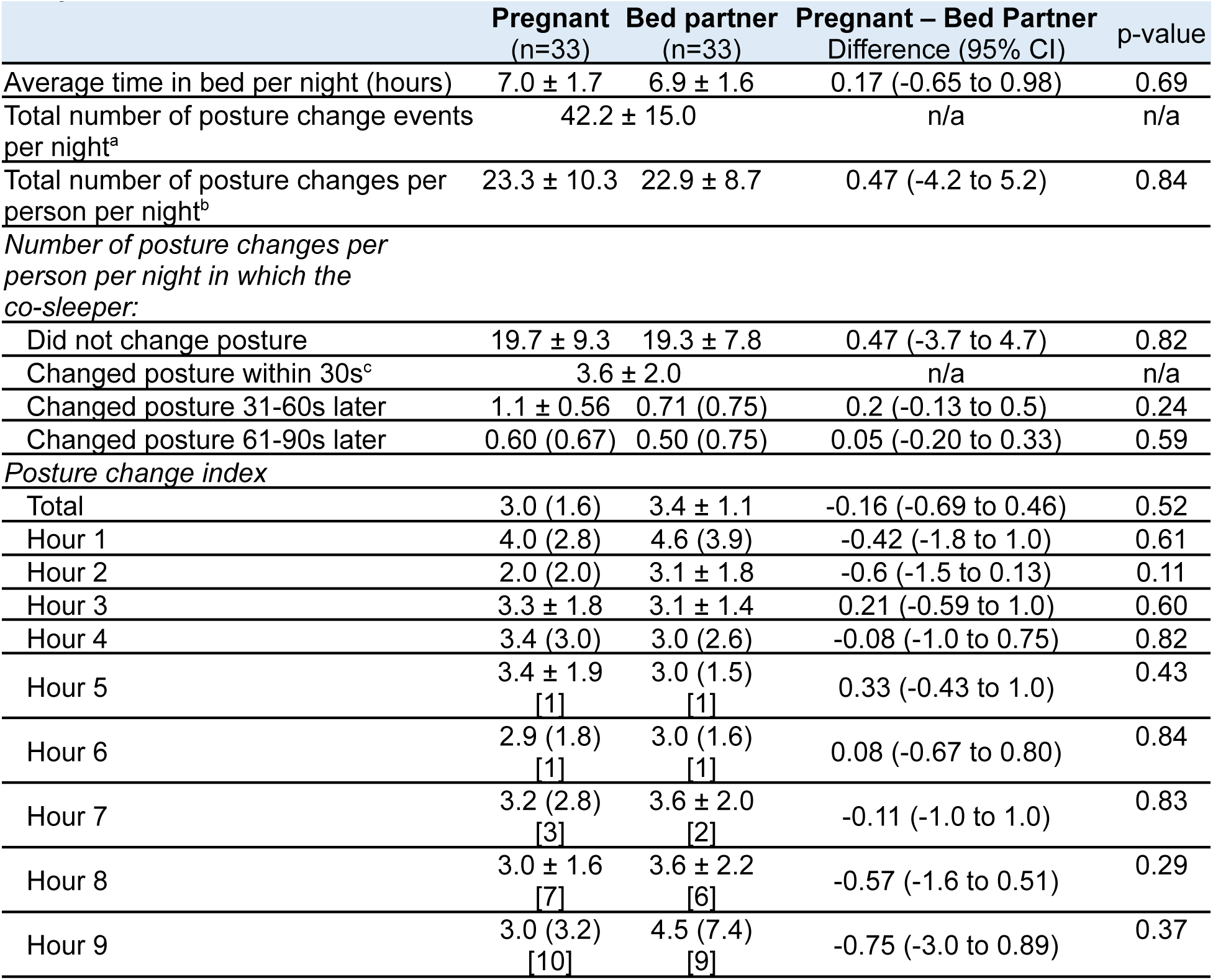
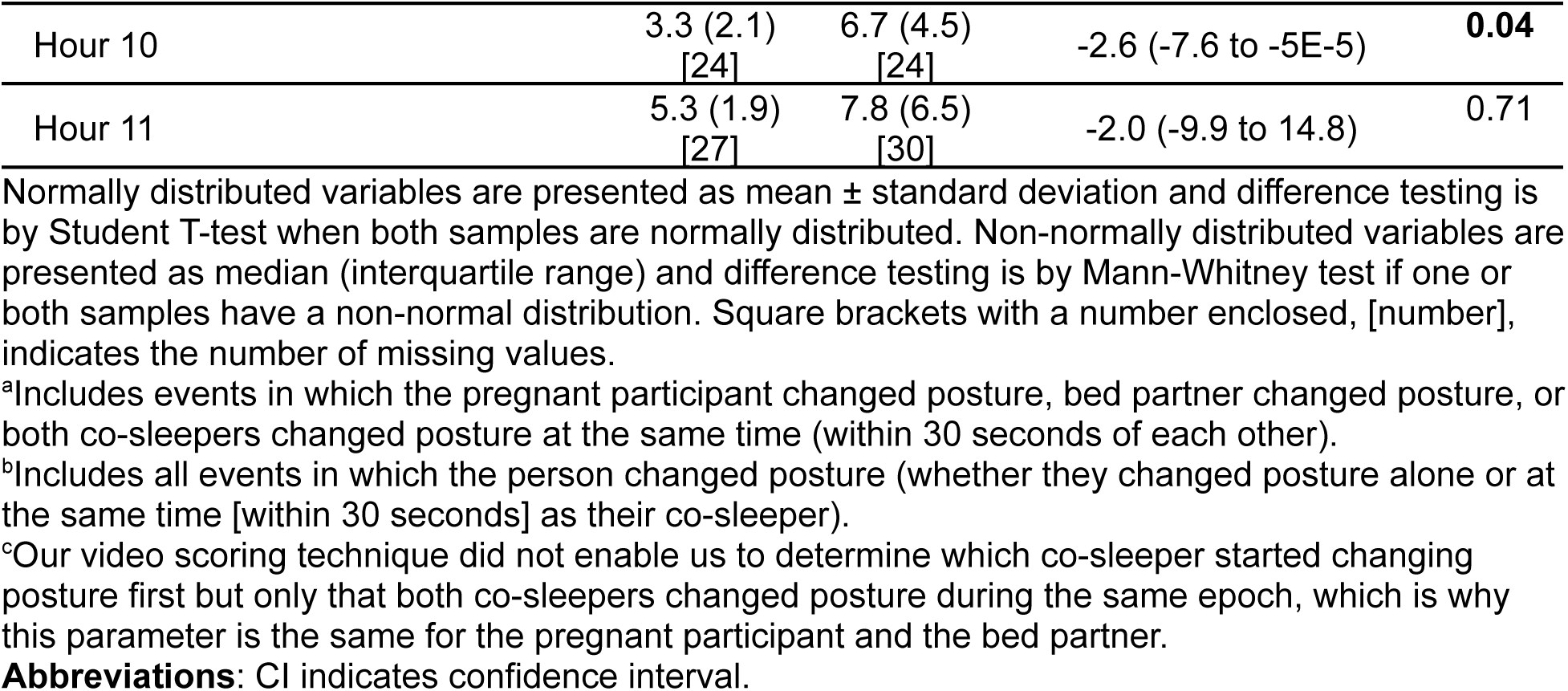
Average time in bed per night, number of posture change events per night, number of posture changes per person per night, posture change interactions, and hour-by-hour posture change index for the pregnant participants and bed partners based on video.

Participants whose bed partner did not participate in the study were excluded from this analysis, but are included in the **Supplementary File**. Furthermore, participants (and their bed partner, if applicable) who completed the sleep studies postpartum were excluded from this analysis and are presented separately in the **Supplementary File**.

For the 33 pregnant participant and bed partner couples, **Figure 2** displays individual mean PCI’s and grouped mean PCI’s as the overnight video recording progressed. Since the individuals had differing lengths of sleep (and, hence, differing lengths of overnight videos; see the increasing number of missing values with increasing hours in **Table 4**), the progression of the video on the x-axis is presented two ways: (1) based on the number of hours of overnight video elapsed (“absolute video length”), and (2) based on the percentage of the overnight video elapsed (“normalized video length”). The normalized plots, therefore, account for differing time in bed and show the general pattern of the PCI as the night progresses, whereas the absolute plots are impacted by decreasing sample size as the night progresses.

**Figure 2.**
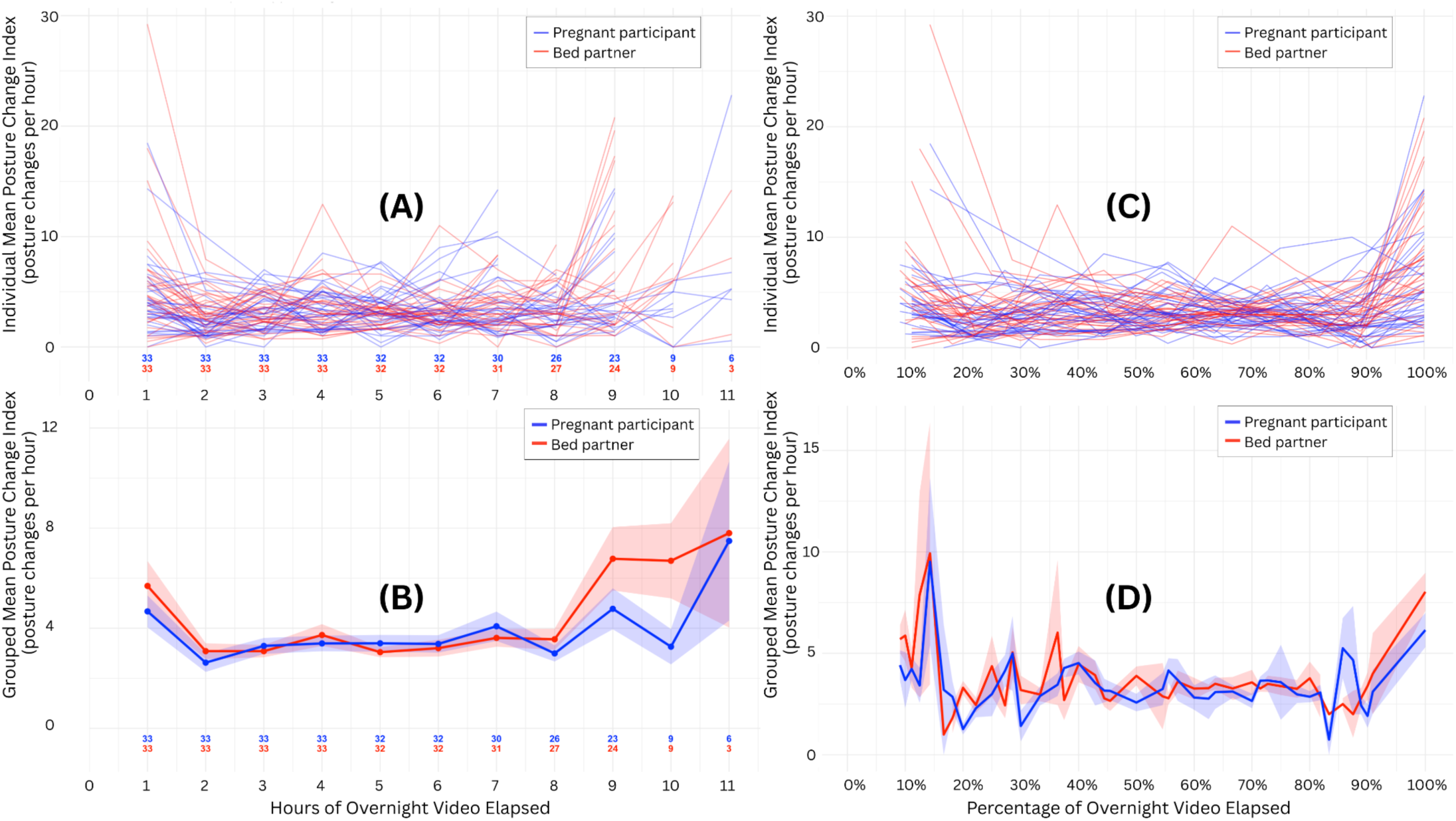
Plots of mean hourly posture change indices for individuals (plots A and C) and pregnant participant and bed partner groups (plots B and D) based on hours (plots A and B) and percentage (plots C and D) of overnight video elapsed for the 33 couples. (A) Mean hourly posture change indices for individuals based on the hours of overnight video elapsed, with pregnant participants represented by blue lines and bed partners represented by red lines. The bold numbers at the bottom of the plot show the sample size remaining at each hour for the pregnant participants (blue font) and bed partners (red font). (B) Mean hourly posture change indices for pregnant participant and bed partner groups based on the hours of overnight video elapsed, with the pregnant participant group represented by the blue line and bed partner group represented by the red line. The points represent the raw mean for each hour, and the shaded area above and below each line represents one standard error of the mean. The bold numbers at the bottom of the plot show the sample size remaining at each hour for the pregnant participants (blue font) and bed partners (red font). (C) Mean hourly posture change indices for individuals based on the percentage of overnight video elapsed, with pregnant participants represented by blue lines and bed partners represented by red lines. (D) Mean hourly posture change indices for pregnant participant and bed partner groups based on the percentage of overnight video elapsed, with the pregnant participant group represented by the blue line and bed partner group represented by the red line. The shaded area above and below each line represents one standard error of the mean.

A Pearson correlation analysis was completed for the 33 pregnant participant and bed partner couples across all hours of all nights (total of 985 hours of matched video data across 135 nights) and showed a statistically significant positive correlation in hourly PCI between pregnant participants and their bed partners (R=0.37; 95%CI 0.31 to 0.42; p<2.2E-16). **Figure 3** displays the results of the PCI correlation analysis graphically. The slope of the line fit by a simple linear regression model is 0.29, which indicates that for every additional posture change that the bed partner makes per hour, the pregnant participant is predicted to make an additional 0.29 posture changes. A simple linear regression was also fit to predict the number of bed partner posture changes based on pregnant participant’s posture changes (not shown), and for every additional posture change that the pregnant participant makes per hour, the bed partner is predicted to make an additional 0.47 posture changes.

**Figure 3.**
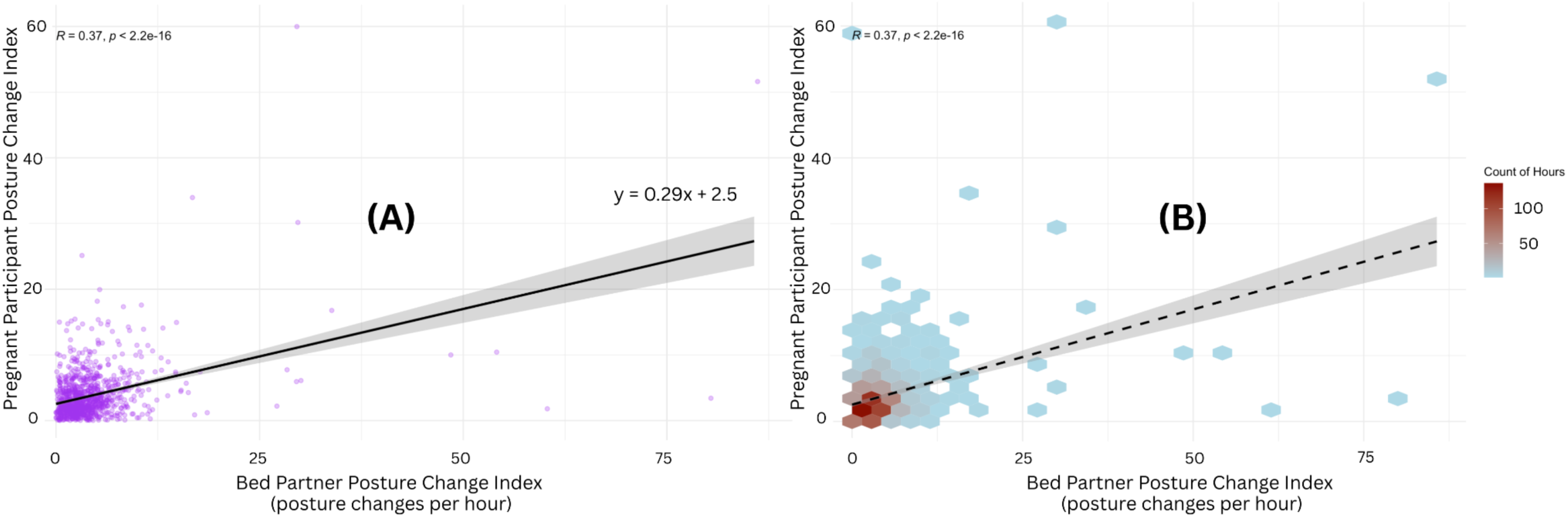
Correlation between pregnant participants’ hourly posture change index (posture changes per hour) and their bed partners’ hourly posture change index (posture changes per hour) for the 33 couples over a total of 985 hours of matched video data across 135 nights. **(A)** Scatter plot of posture change index between each pregnant participant and their bed partner. Each purple dot represents one hour of matched video. Pearson’s R is 0.37, p<2.2E-16, which is a moderately positive correlation, that is, as the pregnant participant’s PCI increases, the bed partner’s PCI increases, and vice versa. **(B)** Two-dimension density plot of the scatter plot in (A) using hexagonal binning. The matched video hours are most concentrated in the bottom left corner, which highlights the peak of synchronous posture changes.

### Sleep Behaviour Characteristics

The cumulative number of absences per night and cumulative time absent from bed per night for the pregnant participants and bed partners based on analysis of the overnight video recordings is shown in **Table 5**. Participants (and their bed partner, if applicable) who completed the sleep studies postpartum were excluded from this analysis and are presented separately in the **Supplementary File**.

**Table 5.** Cumulative number of absences per night and cumulative time (in minutes) spent absent from bed per night for the pregnant participants and bed partners based on video.

|  | Pregnant<br>(n=38) | Bed partner<br>(n=33) | Pregnant – Bed Partner<br>Difference (95% CI) | p-value |
| --- | --- | --- | --- | --- |
| Cumulative number of absences | 1.3 (1.0) | 0.50 (0.80) | 1.0 (0.63 to 1.3) | **<0.001** |
| Cumulative time (minutes) absent | 3.2 (3.7) | 0.88 (1.9) | 2.1 (1.3 to 3.4) | **<0.001** |

Non-normally distributed variables are presented as median (interquartile range) and difference testing is by Mann-Whitney test if one or both samples have a non-normal distribution.
**Abbreviations:** CI indicates confidence interval.

### Impact of Sleep Environment Variables

#### Impact on Cumulative Nightly Time Spent in Each Sleeping Posture

To determine the impact of sleeping environment variables on the cumulative nightly time spent in each sleeping posture, these times were regressed on pregnancy status, the side of the bed the person slept on, and the presence (or absence) of a pregnancy pillow, bed partner, shared bedsheets, and pets. **Table 6** displays the results of a multivariate linear regression of cumulative nightly time spent in each sleeping posture regressed on pregnancy status, the side of the bed the pregnant participant slept on, and presence (or absence) of a pregnancy pillow, bed partner, shared bed sheets, and pets for the pregnant participant.

**Table 6.** Multivariate linear regressions of cumulative nightly time (in minutes) spent in each sleeping posture by pregnant participants regressed on participant’s pregnancy status, the side of the bed the person slept on, and the presence of a pregnancy pillow, a bed partner, shared bed sheets, and pets.

| Posture | Pregnancy status<br>Ref: Postpartum<br>B (SE)<br>95% CI<sup>†</sup> | Side of bed<br>Ref: Right<br>B (SE)<br>95% CI<sup>†</sup> | Pregnancy pillow<br>Ref: Present<br>B (SE)<br>95% CI<sup>†</sup> | Bed partner<br>Ref: Present<br>B (SE)<br>95% CI<sup>†</sup> | Bed sheets<br>Ref: Shared<br>B (SE)<br>95% CI<sup>†</sup> | Pets<br>Ref: Present<br>B (SE)<br>95% CI<sup>†</sup> | R<sup>2</sup> | F |
| --- | --- | --- | --- | --- | --- | --- | --- | --- |
| LR | -14.6 (15.2) | -8.5 (8.7) | -25.4* (9.8)<br>-45.2 to -5.6 | -19.4 (15.8) | 3.2 (11.6) | 8.2 (10.7) | 0.24 | 0.11 |
| LL | -73.0* (33.6)<br>-141.1 to -4.8 | 1.0 (19.2) | 0.2 (21.5) | 38.0 (34.7) | 21.2 (25.5) | 20.1 (23.5) | 0.22 | 0.17 |
| LT | -15.1 (15.0) | -14.1 (8.6) | 1.6 (9.6) | -10.3 (15.5) | 15.8 (11.4) | 25.4* (10.5)<br>4.0 to 46.7 | 0.26 | 0.09 |
| S | 152.0*** (31.9)<br>87.3 to 216.8 | -18.0 (18.2) | 22.5 (20.4) | 39.8 (33.0) | 0.64 (24.2) | 35.5 (22.3) | 0.45 | .001 |
| STRP | -5.1 (7.1) | 1.9 (4.1) | -3.4 (4.5) | 3.8 (7.3) | 4.0 (5.4) | 1.7 (5.0) | 0.08 | 0.81 |
| STLP | 11.0 (6.2) | -4.5 (3.5) | 0.56 (4.0) | 4.2 (6.4) | 1.2 (4.7) | 3.3 (4.3) | 0.15 | 0.41 |
| SPRT | 0.096 (1.0) | 0.63 (0.58) | 0.47 (0.65) | 0.29 (1.0) | 0.64 (0.76) | 1.3 (0.70) | 0.18 | 0.30 |
| SPLT | -0.45 (1.6) | 0.45 (0.94) | 0.99 (1.0) | 3.3 (1.7) | -2.7* (1.2)<br>-5.2 to -0.14 | 0.6 (1.1) | 0.23 | 0.15 |
| RT | -3.3 (21.2) | 10.1 (12.1) | 21.0 (13.6) | -46.0* (21.9)<br>-90.4 to -1.5 | 2.8 (16.1) | -17.5 (14.8) | 0.24 | 0.11 |
| RL | -56.3 (40.4) | 2.9 (23.1) | -27.4 (25.9) | -66.9 (41.8) | 94.5** (30.7)<br>32.2 to 156.8 | 9.5 (28.3) | 0.30 | 0.05 |
| RR | -4.2 (16.3) | 20.3* (9.3)<br>1.4 to 39.2 | -7.8 (10.4) | -49.4** (16.8)<br>-83.5 to -15.3 | 5.7 (12.3) | 5.0 (11.4) | 0.33 | 0.02 |
| P | n/a | n/a | n/a | n/a | n/a | n/a | n/a | n/a |
| Sit | 15.3** (5.1)<br>5.0 to 25.6 | -3.5 (2.9) | 0.65 (3.2) | -2.5 (5.2) | 3.1 (3.8) | 0.42 (3.5) | 0.25 | 0.10 |

\*p<0.05, \*\*p<0.01, \*\*\*p<0.001
<sup>†</sup>The 95% CI is given if the coefficient is statistically significant.
**Abbreviations:** B indicates the coefficient of the variable; SE indicates the standard error of the coefficient; CI indicates confidence interval; R<sup>2</sup> indicates the R-squared value of the explanatory model; F indicates the F-statistic of the explanatory model; LR indicates left recovery; LL indicates left lateral; LT indicates left tilt; S indicates supine; STRP indicates supine thorax with right pelvic tilt; STLP indicates supine thorax with left pelvic tilt; SPRT indicates supine pelvis with right thorax tilt; SPLT indicates supine pelvis with left thorax tilt; RT indicates right tilt; RL indicates right lateral; RR indicates right recovery; P indicates prone; Sit indicates sitting.

For the bed partners, cumulative nightly time spent in each sleeping posture was regressed on the pregnancy status of their partner (i.e., the pregnant participant), the side of the bed that the bed partner slept on, and the presence (or absence) of a pregnancy pillow, shared bed sheets, and pets (see **Table 7**). The pregnancy status of the pregnant participant and presence or absence of shared bed sheets did not have a statistically significant effect on the bed partner’s cumulative nightly time spent in a given sleeping posture in any of the multivariate models. Bed partners who slept on the right side of the bed spent 31.8 less minutes in left lateral (p 0.053), 13.6 less minutes in left tilt, 47.9 more minutes in right lateral, and 29.2 more minutes in right recovery. In the presence of a pregnancy pillow, bed partners spent 0.95 more minutes sitting. In the presence of pets, bed partners spent 50.9 more minutes in the right lateral posture.

**Table 7.**
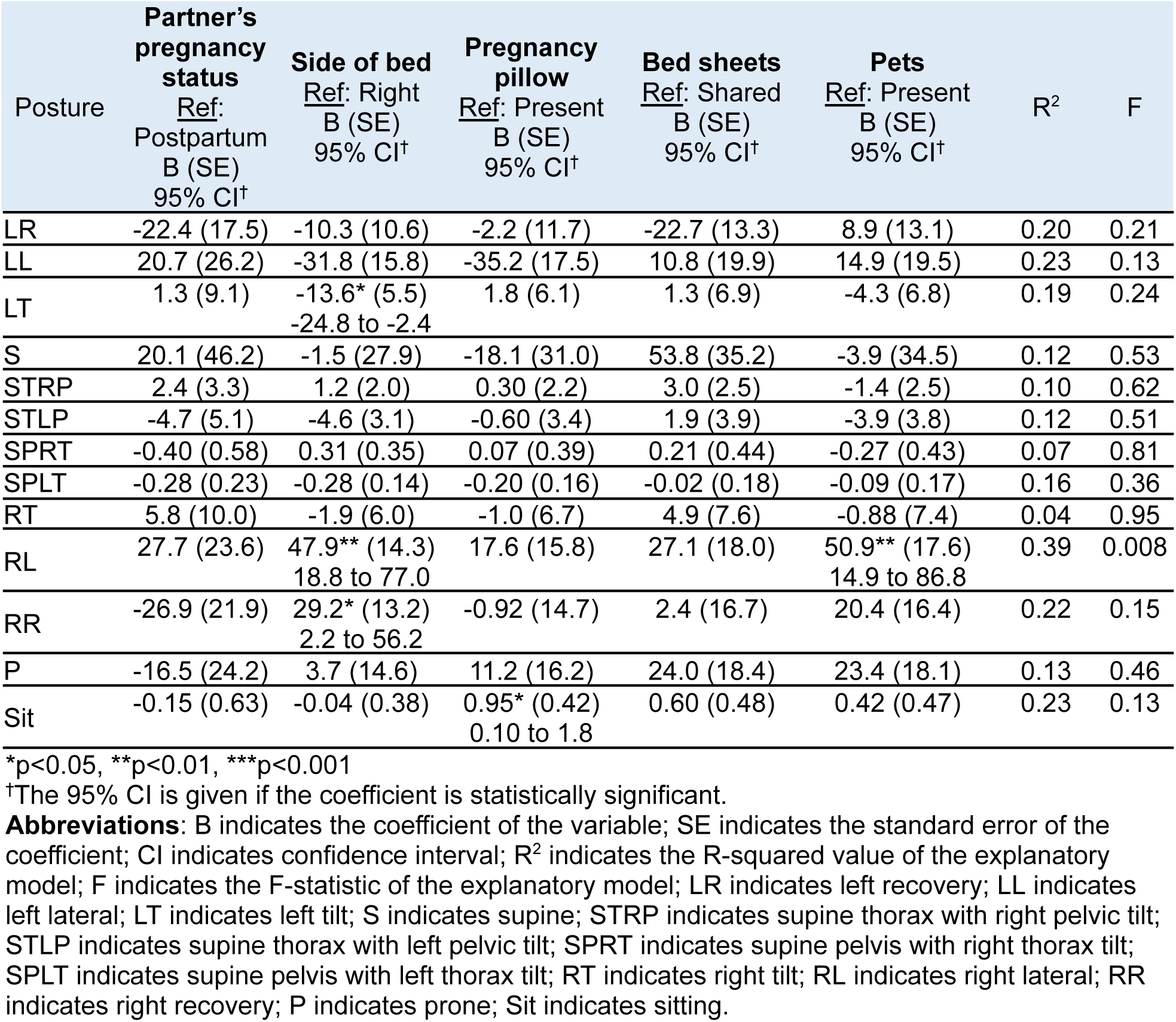
Multivariate linear regressions of cumulative nightly time (in minutes) spent in each sleeping posture by bed partners regressed on their partner’s pregnancy status, the side of the bed the person slept on, and the presence of a pregnancy pillow, shared bed sheets, and pets.

#### Impact on Episodic Time Spent in Each Sleeping Posture

To determine the impact of sleeping environment variables on episodic time spent in each sleeping posture, these times were regressed on pregnancy status, the side of the bed the person slept on, and the presence (or absence) of a pregnancy pillow, bed partner, shared bedsheets, and pets. **Table 8** shows the average duration of episodes that the pregnant participant spent in each sleeping posture regressed on sleeping environment variables. The side of the bed the pregnant participant slept on and the presence or absence of a pregnancy pillow, bed partner, and shared bed sheets did not have a statistically significant effect on the average duration of sleeping posture episodes in any of the multivariate models. Pregnant participants who underwent the study in the antepartum period had a shorter average duration of episodes in the supine thorax with left pelvic tilt and sitting postures (21.2 and 8.3 minutes shorter, respectively). Pregnant participants with pets present had shorter average duration of episodes (13 minutes shorter) in the left lateral posture.

**Table 8.** Multivariate linear regressions of the average duration (in minutes) of episodes spent in each sleeping posture by pregnant participants regressed on participant’s pregnancy status, the side of the bed the participant slept on, and the presence of a pregnancy pillow, bed partner, shared bed sheets, and pets.

| Posture | Pregnancy status<br>Ref: Postpartum<br>B (SE)<br>95% CI† | Side of bed<br>Ref: Right<br>B (SE)<br>95% CI† | Pregnancy pillow<br>Ref: Present<br>B (SE)<br>95% CI† | Bed partner<br>Ref: Present<br>B (SE)<br>95% CI† | Bed sheets<br>Ref: Shared<br>B (SE)<br>95% CI† | Pets<br>Ref: Present<br>B (SE)<br>95% CI† | R<sup>2</sup> | F |
| --- | --- | --- | --- | --- | --- | --- | --- | --- |
| LR | -3.7 (10.6) | -4.8 (7.0) | -8.7 (8.3) | -9.1 (12.7) | 11.3 (11.1) | -3.9 (9.2) | 0.25 | 0.96 |
| LL | -12.7 (7.7) | 6.4 (4.5) | 3.6 (5.0) | 11.6 (8.6) | -0.81 (5.9) | -13.0* (5.6)<br>-24.3 to -1.8 | 0.27 | 2.1 |
| LT | -7.6 (7.0) | -3.7 (3.5) | -1.4 (3.6) | -1.2 (5.8) | 5.0 (4.6) | -1.2 (3.7) | 0.19 | 1.1 |
| S | 15.9 (7.8) | -5.6 (4.7) | 3.8 (5.4) | 9.0 (8.2) | 0.79 (6.4) | 0.23 (5.4) | 0.21 | 1.4 |
| STRP | 2.4 (10.4) | 3.3 (5.2) | 2.9 (5.6) | 8.6 (9.9) | -3.4 (7.4) | -7.3 (6.5) | 0.12 | 0.47 |
| STLP | 21.2* (7.5)<br>5.4 to 37.0 | -1.7 (4.2) | 0.87 (4.4) | -1.2 (6.9) | 9.0 (5.1) | 2.5 (4.7) | 0.41 | 2.1 |
| SPRT | n/a | n/a | n/a | n/a | n/a | n/a | n/a | n/a |
| SPLT | n/a | n/a | n/a | n/a | n/a | n/a | n/a | n/a |
| RT | 0.04 (13.9) | -4.6 (5.2) | 6.4 (5.2) | -1.5 (8.7) | -4.6 (7.8) | -9.6 (6.1) | 0.13 | 0.64 |
| RL | -11.7 (8.6) | 1.3 (4.9) | -1.8 (5.5) | -7.1 (8.8) | 10.9 (6.6) | -12.2 (6.2) | 0.26 | 2.0 |
| RR | 3.8 (7.5) | -4.0 (5.0) | -3.1 (5.7) | -5.3 (9.8) | 7.8 (9.6) | 0.98 (7.1) | 0.19 | 0.72 |

| P | n/a | n/a | n/a | n/a | n/a | n/a | n/a | n/a |
| --- | --- | --- | --- | --- | --- | --- | --- | --- |
| Sit | 8.3** (2.5)<br>3.2 to 13.5 | -0.40 (1.4) | -0.80 (1.4) | -0.79 (2.6) | 2.2 (1.5) | 0.18 (1.4) | 0.43 | 2.8 |

\*p<0.05, \*\*p<0.01, \*\*\*p<0.001
†The 95% CI is given if the coefficient is statistically significant.
**Abbreviations:** B indicates the coefficient of the variable; SE indicates the standard error of the coefficient; CI indicates confidence interval; R<sup>2</sup> indicates the R-squared value of the explanatory model; F indicates the F-statistic of the explanatory model; LR indicates left recovery; LL indicates left lateral; LT indicates left tilt; S indicates supine; STRP indicates supine thorax with right pelvic tilt; STLP indicates supine thorax with left pelvic tilt; SPRT indicates supine pelvis with right thorax tilt; SPLT indicates supine pelvis with left thorax tilt; RT indicates right tilt; RL indicates right lateral; RR indicates right recovery; P indicates prone; Sit indicates sitting.

**Table 9** shows the maximum duration of episodes that the pregnant participant spent in each sleeping posture regressed on sleeping environment variables. The side of the bed the pregnant participant slept on and the presence or absence of a pregnancy pillow and pet did not have a statistically significant effect on the maximum duration of sleeping posture episodes in any of the multivariate models. Pregnant participants who underwent the study in the antepartum period had a shorter maximum duration of episodes in the supine and supine thorax with left pelvic tilt postures (75.8 and 32.8 minutes shorter, respectively). Pregnant participants who shared bed sheets with their bed partner had a longer maximum duration of episodes in the right lateral posture (49.4 minutes longer).

**Table 9.**
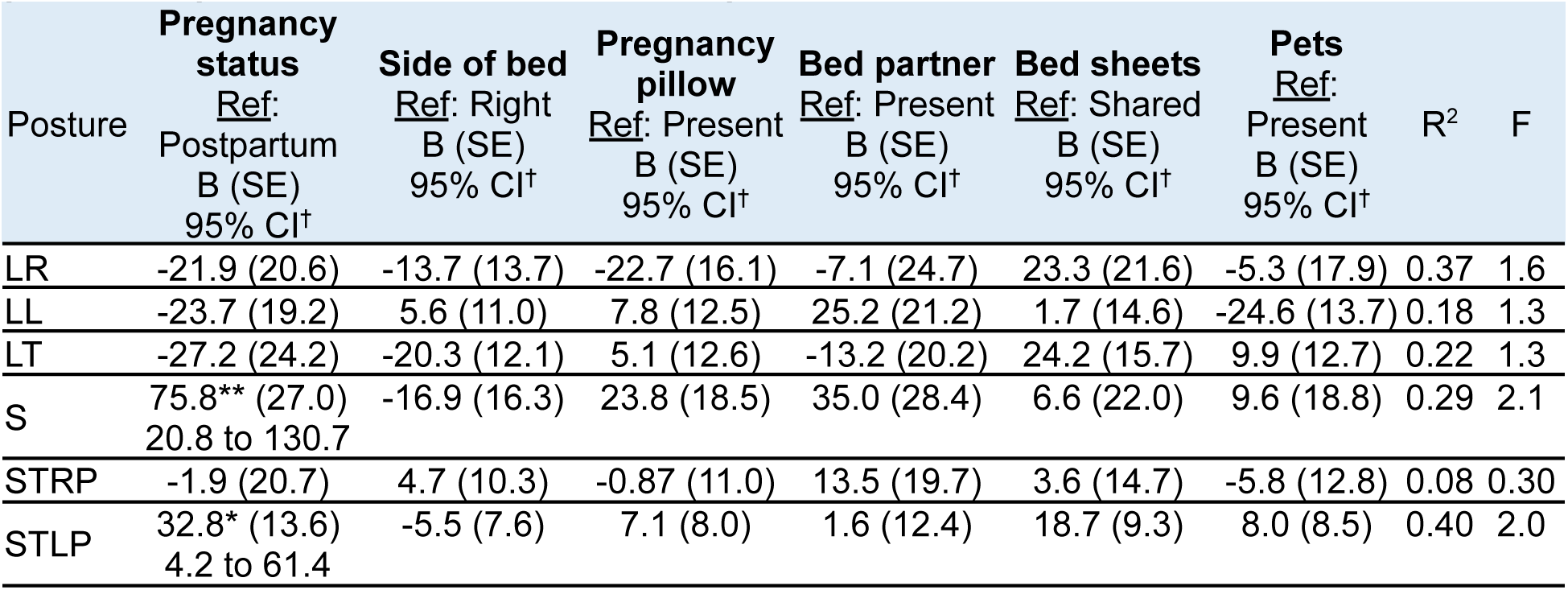
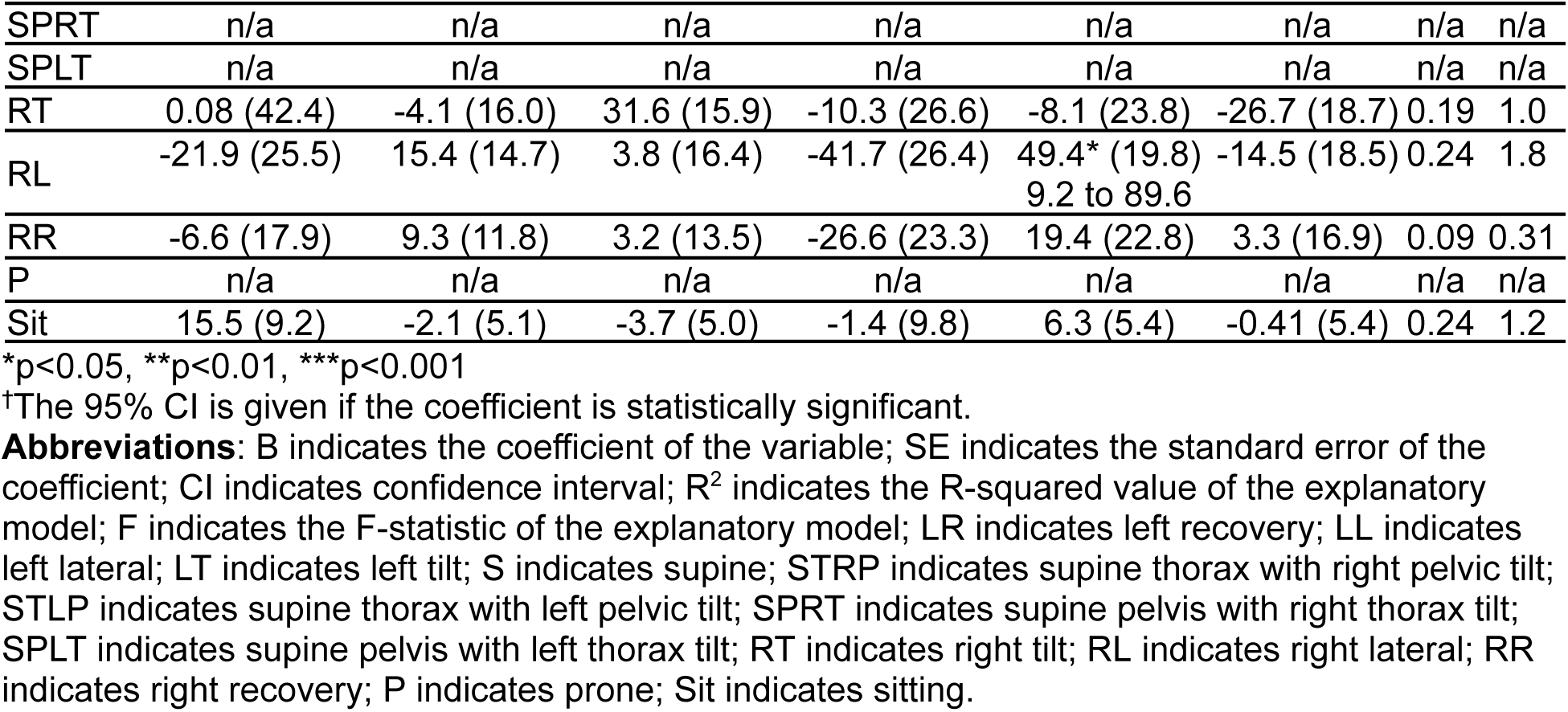
Multivariate linear regressions of the maximum duration (in minutes) of episodes spent in each sleeping posture by pregnant participants regressed on participant’s pregnancy status, side of the bed the participant sleeps on, and presence of a pregnancy pillow, bed partner, shared bedsheets, and pets.

**Table 10** shows the average duration of episodes that the bed partner spent in each sleeping posture regressed on sleeping environment variables. The side of the bed the bed partner slept on did not have a statistically significant effect on the bed partner’s average duration of sleeping posture episodes in any of the multivariate models. Bed partners who underwent the study when their partner was in the postpartum period had a longer average duration of episodes (by 11.6 minutes) in the left lateral posture. In the presence of a pregnancy pillow, bed partners had a longer average duration of episodes in the prone posture (by 13.4 minutes). Bed partners who shared bed sheets with their partner had a longer average duration of episodes in the supine thorax with right pelvic tilt posture (by 6.5 minutes).

**Table 10.**
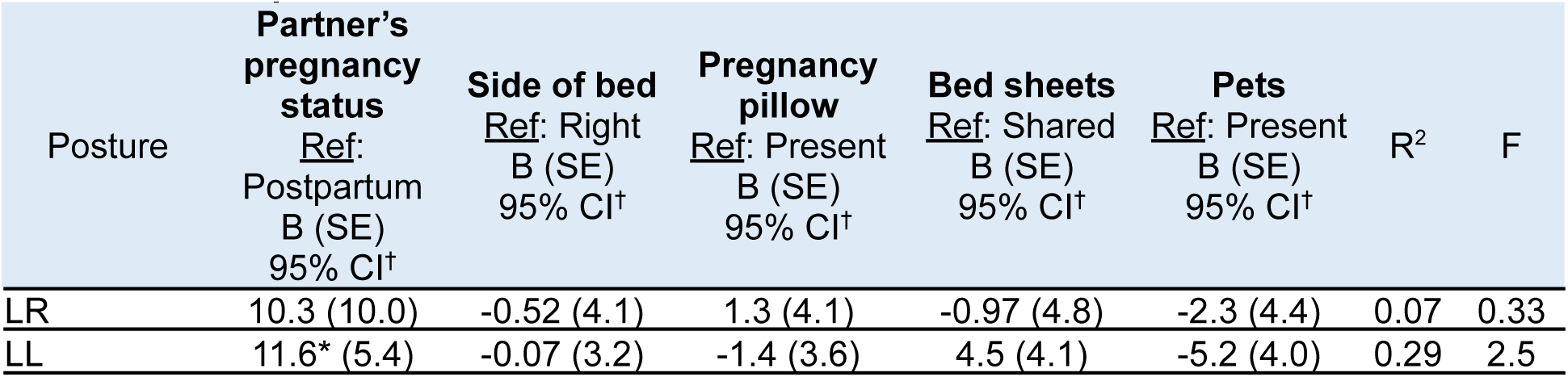
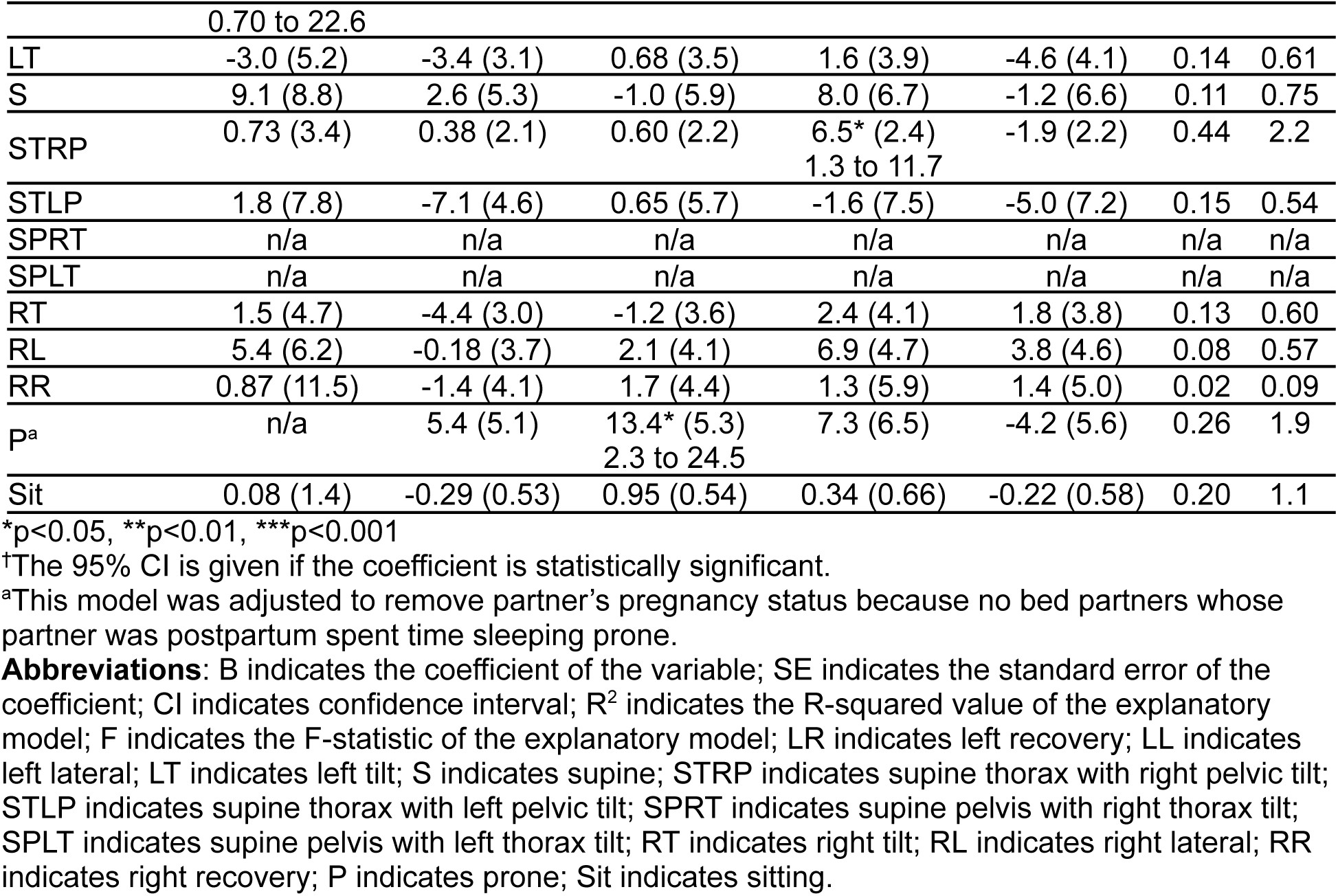
Multivariate linear regressions of the average duration (in minutes) of episodes spent in each sleeping posture by bed partners regressed on their partner’s pregnancy status, the side of the bed they slept on, and the presence of a pregnancy pillow, shared bedsheets, and pets.

**Table 11** shows the maximum duration of episodes that the bed partner spent in each sleeping posture regressed on sleeping environment variables. The pregnancy status of the pregnant participant did not have a statistically significant effect on the bed partner’s maximum duration of sleeping posture episodes in any of the multivariate models. In the presence of a pregnancy pillow, bed partners had a longer maximum duration of episodes in the prone posture (by 28.7 minutes). Bed partners who slept on the right side of the bed had shorter maximum duration of episodes in the left tilt posture (20.2 minutes shorter) and longer maximum duration of episodes in the right lateral posture (19.6 minutes longer). Bed partners who shared bed sheets with their partner had a longer maximum duration of episodes in the supine thorax with right pelvic tilt posture (by 19.3 minutes) and right lateral posture (by 21.3 minutes). Bed partners with pets present had a longer average duration of episodes in the right lateral posture (by 20.1 minutes).

**Table 11.** Multivariate linear regressions of the maximum duration (in minutes) of episodes spent in each sleeping posture by bed partners regressed on their partner’s pregnancy status, presence of a pregnancy pillow, side of the bed they sleep on, shared or separate bedsheets, and presence of pets.

| Posture | Partner's pregnancy status<br>Ref: Postpartum<br>B (SE)<br>95% CI<sup>†</sup> | Side of bed<br>Ref: Right<br>B (SE)<br>95% CI<sup>†</sup> | Pregnancy pillow<br>Ref: Present<br>B (SE)<br>95% CI<sup>†</sup> | Bed sheets<br>Ref: Shared<br>B (SE)<br>95% CI<sup>†</sup> | Pets<br>Ref: Present<br>B (SE)<br>95% CI<sup>†</sup> | R<sup>2</sup> | F |
| --- | --- | --- | --- | --- | --- | --- | --- |
| LR | 33.8 (23.4) | 0.94 (9.5) | 10.8 (9.7) | -11.5 (11.2) | 0.09 (10.2) | 0.19 | 1.0 |
| LL | 19.8 (14.1) | -4.4 (8.5) | -4.1 (9.4) | 9.9 (10.7) | -4.1 (10.5) | 0.14 | 1.0 |
| LT | -2.6 (14.6) | -20.2* (8.8)<br>-38.7 to -1.7 | 6.8 (9.9) | 5.1 (11.0) | -9.4 (11.6) | 0.25 | 1.3 |
| S | 14.8 (33.8) | 11.9 (20.4) | -2.1 (22.6) | 35.5 (25.7) | 7.3 (25.2) | 0.08 | 0.53 |
| STRP | 18.9 (11.9) | -9.1 (7.6) | 4.9 (8.0) | 19.3* (8.6)<br>0.81 to 37.7 | -7.5 (7.8) | 0.44 | 2.2 |
| STLP | -12.4 (16.6) | -15.9 (9.7) | 0.94 (12.1) | 3.4 (15.9) | -15.9 (15.4) | 0.21 | 0.77 |
| SPRT | n/a | n/a | n/a | n/a | n/a | n/a | n/a |
| SPLT | n/a | n/a | n/a | n/a | n/a | n/a | n/a |
| RT | 2.4 (13.8) | -1.9 (8.9) | 4.6 (10.7) | 15.8 (12.1) | 6.2 (11.3) | 0.08 | 0.36 |
| RL | 7.7 (12.9) | 19.6* (7.8)<br>3.8 to 35.5 | 8.8 (8.6) | 21.3* (9.8)<br>1.3 to 41.3 | 20.1* (9.6)<br>0.51 to 39.8 | 0.30 | 2.6 |
| RR | 5.3 (28.3) | 6.2 (10.2) | 8.2 (11.0) | 15.1 (14.5) | 21.8 (12.3) | 0.14 | 0.79 |
| P<sup>a</sup> | n/a | 10.6 (11.7) | 28.7* (12.2)<br>3.2 to 54.1 | 30.8 (14.9) | 1.9 (12.8) | 0.27 | 1.9 |
| Sit | -0.13 (2.6) | 0.15 (1.0) | 1.8 (1.0) | 0.79 (1.2) | -0.03 (1.1) | 0.15 | 0.75 |

\*p<0.05, \*\*p<0.01, \*\*\*p<0.001
<sup>†</sup>The 95% CI is given if the coefficient is statistically significant.
<sup>a</sup>This model was adjusted to remove partner's pregnancy status because no bed partners whose partner was postpartum spent time sleeping prone.
**Abbreviations:** B indicates the coefficient of the variable; SE indicates the standard error of the coefficient; CI indicates confidence interval; R<sup>2</sup> indicates the R-squared value of the explanatory model; F indicates the F-statistic of the explanatory model; LR indicates left recovery; LL indicates left lateral; LT indicates left tilt; S indicates supine; STRP indicates supine thorax with right pelvic tilt; STLP indicates supine thorax with left pelvic tilt; SPRT indicates supine pelvis with right thorax tilt; SPLT indicates supine pelvis with left thorax tilt; RT indicates right tilt; RL indicates right lateral; RR indicates right recovery; P indicates prone; Sit indicates sitting.

#### Impact on Sleeping Posture Changes

A multivariate linear regression of the pregnant participants’ and bed partners’ total number of posture changes per night regressed on pregnancy status, the side of the person slept on, and the presence (or absence) of a pregnancy pillow, bed partner, shared bedsheets, child/children, and pet/pets (see **Table 12**). In the presence of pets, both pregnant participants and bed partners experienced a significant increase in the total number of posture changes per night (17.3 and 8.0, respectively) when compared to in the absence of pets.

**Table 12.** Multivariate linear regression of a person’s total number of posture changes per night regressed on the pregnant participant’s pregnancy status, side of the bed the person sleeps on, and the presence of a pregnancy pillow, a bed partner, shared bedsheets, a child/children, and a pet/pets.

|  | Pregnant Participant |  |  |  | Bed Partner |  |  |  |
| --- | --- | --- | --- | --- | --- | --- | --- | --- |
|  | B | SE | 95% CI | p-value | B | SE | 95% CI | p-value |
| Pregnancy status<br>Ref: Postpartum | -3.6 | 4.1 | -12.0 to 4.9 | 0.40 | -5.6 | 4.7 | -15.2 to 4.0 | 0.24 |
| Side of bed<br>Ref: Right | -3.4 | 2.4 | -8.2 to 1.5 | 0.17 | 3.0 | 2.9 | -2.9 to 8.9 | 0.30 |
| *Presence of:* |  |  |  |  |  |  |  |  |
| Pregnancy pillow<br>Ref: Yes | -3.3 | 2.6 | -8.7 to 2.0 | 0.22 | -2.4 | 3.1 | -8.8 to 4.0 | 0.45 |
| Bed partner<br>Ref: Yes | -7.7 | 4.4 | -16.8 to 1.3 | 0.09 | n/a | n/a | n/a | n/a |
| Shared bedsheets<br>Ref: Yes | 5.5 | 3.1 | -0.90 to 11.9 | 0.09 | 0.53 | 3.6 | -6.8 to 7.8 | 0.88 |
| Child/children<br>Ref: Yes | 1.6 | 3.5 | -5.5 to 8.7 | 0.654 | 5.5 | 4.7 | -4.1 to 15.1 | 0.25 |
| Pet/pets<br>Ref: Yes | 17.3 | 2.9 | 11.5 to 23.2 | **< .001** | 8.0 | 3.6 | 0.74 to 15.3 | **0.03** |
| R<sup>2</sup>=0.56, F=6.3, p<0.001 |  |  |  |  | R<sup>2</sup>=0.24, F=1.5, p=0.20 |  |  |  |

**Abbreviations:** B indicates the coefficient of the variable; SE indicates the standard error of the coefficient; CI indicates confidence interval; R<sup>2</sup> indicates the R-squared value of the explanatory model; F indicates the F-statistic of the explanatory model.

#### Impact on Sleeping Behaviour

A multivariate linear regression of the pregnant participants’ and bed partners’ cumulative nightly time absent from bed was regressed on pregnancy status, the side of the person slept on, and the presence (or absence) of a pregnancy pillow, bed partner, shared bedsheets, child/children, and pet/pets (see **Table 13**). Bed partners with pets present spent more time (2.8 minutes) absent from bed per night than those without pets. This phenomenon was also observed in the pregnant participants but did not reach statistical significance.

**Table 13.**
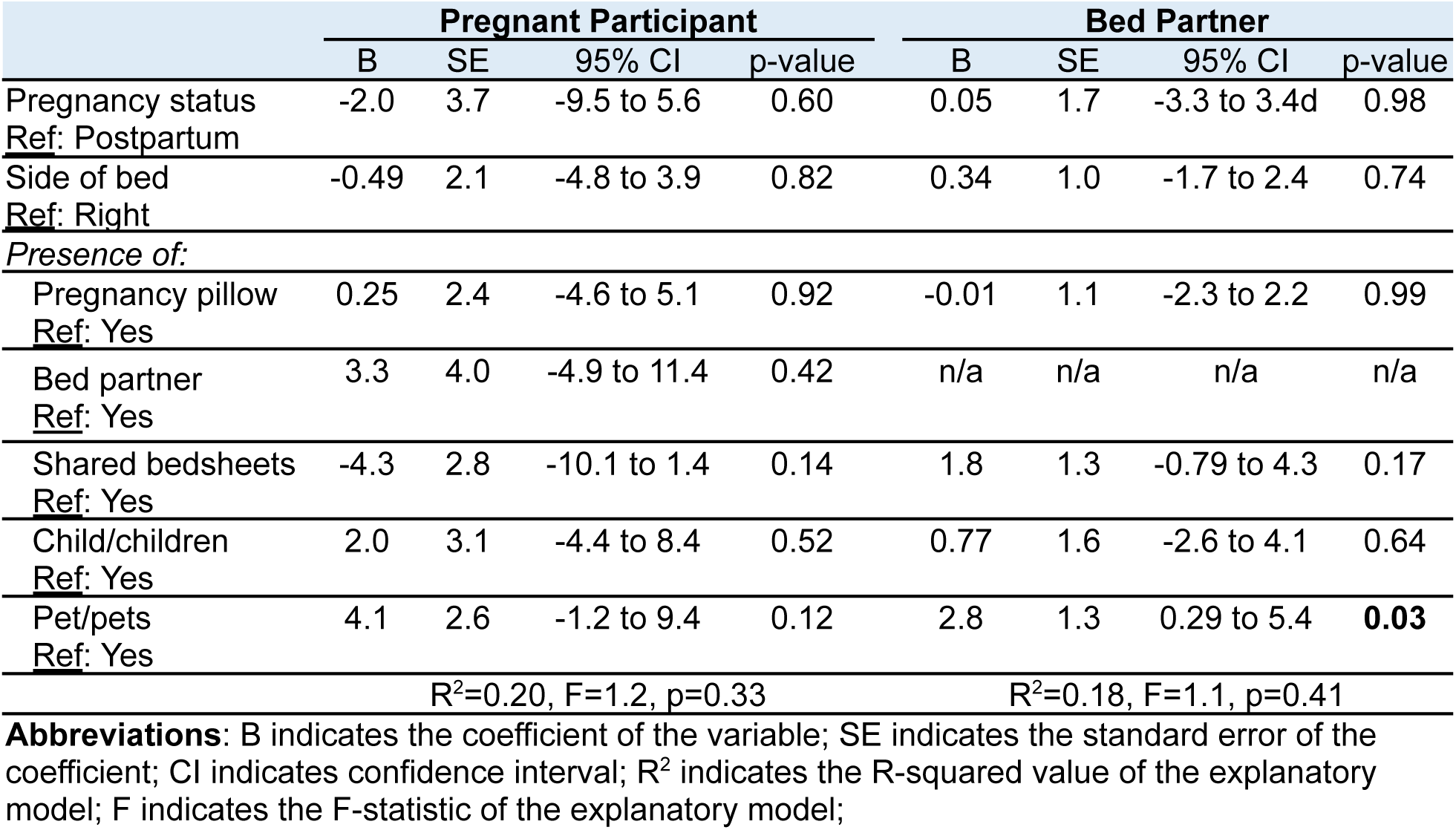
Multivariate linear regressions of a person’s cumulative nightly time (in minutes) absent from bed regressed on the pregnant participant’s pregnancy status, side of bed the person sleeps on, and the presence of a pregnancy pillow, a bed partner, shared bedsheets, a child/children, and a pet/pets.

|  | Pregnant Participant |  |  |  | Bed Partner |  |  |  |
| --- | --- | --- | --- | --- | --- | --- | --- | --- |
|  | B | SE | 95% CI | p-value | B | SE | 95% CI | p-value |
| Pregnancy status<br>Ref: Postpartum | -2.0 | 3.7 | -9.5 to 5.6 | 0.60 | 0.05 | 1.7 | -3.3 to 3.4d | 0.98 |
| Side of bed<br>Ref: Right | -0.49 | 2.1 | -4.8 to 3.9 | 0.82 | 0.34 | 1.0 | -1.7 to 2.4 | 0.74 |
| *Presence of:* |  |  |  |  |  |  |  |  |
| Pregnancy pillow<br>Ref: Yes | 0.25 | 2.4 | -4.6 to 5.1 | 0.92 | -0.01 | 1.1 | -2.3 to 2.2 | 0.99 |
| Bed partner<br>Ref: Yes | 3.3 | 4.0 | -4.9 to 11.4 | 0.42 | n/a | n/a | n/a | n/a |
| Shared bedsheets<br>Ref: Yes | -4.3 | 2.8 | -10.1 to 1.4 | 0.14 | 1.8 | 1.3 | -0.79 to 4.3 | 0.17 |
| Child/children<br>Ref: Yes | 2.0 | 3.1 | -4.4 to 8.4 | 0.52 | 0.77 | 1.6 | -2.6 to 4.1 | 0.64 |
| Pet/pets<br>Ref: Yes | 4.1 | 2.6 | -1.2 to 9.4 | 0.12 | 2.8 | 1.3 | 0.29 to 5.4 | **0.03** |
| R<sup>2</sup>=0.20, F=1.2, p=0.33 |  |  |  |  | R<sup>2</sup>=0.18, F=1.1, p=0.41 |  |  |  |

**Abbreviations:** B indicates the coefficient of the variable; SE indicates the standard error of the coefficient; CI indicates confidence interval; R<sup>2</sup> indicates the R-squared value of the explanatory model; F indicates the F-statistic of the explanatory model;

## Discussion

### Cumulative and Episodic Time Spent in Sleeping Postures

Novel findings from this study show that pregnant participants’ cumulative time spent in various sleeping postures followed a remarkably different pattern from their bed partners and opens a discussion about the impact of pregnancy-related constraints and the sleeping environment on sleep during pregnancy. As expected, pregnant participants did not sleep prone, spent significantly less time in left recovery, right recovery, and supine postures, and spent significantly more time in left lateral, left tilt, and right lateral postures. In addition to the supine posture being uncomfortable in late pregnancy,^58–61^ another possible explanation for more lateral and less supine time could be increased awareness of safe going to sleep posture recommendations among pregnant people and their healthcare providers,^62–66^ but we did not assess this level of awareness. In our study, the median (interquartile range; IQR) cumulative percentage of the night spent supine by the pregnant participants was 3.8% (12.9), which is significantly lower than most reports using objective measures (i.e., position sensor-based or video-determined) in the third trimester. In sleep lab-based studies, means of 15 to 25% (Maasilta et al.; sensor)^45^ and 19.1% (Wilson et al.; sensor)^46^ and medians of 16.4% (Kember et al.; video)^50^ and 26.3% (Wilson et al.; sensor and video)^35^ have been reported. Whereas in home-based studies, means of 16.8% (Warland & Dorian; sensor)^48^ and 47% (Dunietz et al.; sensor)^51^ and medians of 1.1% (Lucchini et al.; sensor),^67^ 9.0% (Wilson et al.; sensor),^36^ 9.5% (Warland et al.; video),^34^ 16.8% (Stone et al.; video),^68^ 19% (McIntyre et al.; video),^49^ 19.0% (Wilson et al.; sensor),^35^ and 26.5% (O’Brien & Warland; sensor)^47^ have been reported. All these studies except Warland & Dorian (mean 16.8%; three nights)^48^ and Wilson et al. (median 9.0%; seven consecutive nights)^36^ were single-night studies and may, therefore, be impacted by a “first night effect”, which could skew the percentage of time supine in either direction. Furthermore, all these studies used a five-posture classification system (e.g., left, right, prone, supine, upright), so a lower percentage of the night spent supine in our sample may be explained by our thirteen-posture classification system, which discriminated between supine and six near-supine postures (i.e., left tilt, supine thorax with left pelvic tilt, supine thorax with right pelvic tilt, supine pelvis with left thorax tilt, supine pelvis with right thorax tilt, and right tilt) that a five-posture classification would simply categorize as “supine”. In fact, if the median percentages of the night spent in each of these six near-supine postures in our sample are combined with the supine time, the result is a median 7.9%, which is comparable to the median of 9.0% reported in Wilson et al.’s seven-night, sensor-based, in-home study in n=32 pregnant participants in the third trimester.^36^

Less cumulative nightly time spent in left and right recovery postures by pregnant participants may be due to the difficulty assuming these postures as a result of obstruction by the ventrally-located gravid uterus. Related to this, on our adjusted analysis, we found that the presence of a pregnancy pillow was associated with 25.4 less minutes per night spent in the left recovery posture (statistically significant; SS), which we suggest is a result of the pregnancy pillow itself, which typically wraps from the back, between the legs, to front of the body in a U-shape, providing an obstruction to the upper (right) leg and thorax from rotating toward the sleeping surface. We also observed that 7.8 less minutes per night were spent in the right recovery posture in the presence of a pregnancy pillow, but this was not statistically significant (NSS). A similar effect was seen in the presence of a bed partner (i.e., less time spent in recovery postures) with 49.4 less minutes per night spent in the right recovery posture (SS) and 19.4 less minutes per night spent in the left recovery posture (NSS), which may be explained by the bed partner providing an obstruction to assuming the recovery postures. A potentially confounding factor that we did not measure nor adjust for was the size of the sleeping surface (i.e., double-, queen-, king-sized mattress), which warrants further investigation.

Not one of the pregnant participants in our sample assumed the prone posture at any time during the four nights, which supports the idea that the gravid uterus may present an obstruction to assuming near-prone postures (i.e., recovery postures) in the third trimester. The complete absence of time spent prone in our pregnant participants generally agrees with all studies of sleeping posture in late pregnancy to date, where prone time is reported as zero or minimal,^35,36,47,48,50,67^ with only one contradictory report (in-home; single-night; not video-based; posture sensor was located on the sternum).^51^ Compression of the maternal retroperitoneal vasculature between the gravid uterus and maternal skeleton could be worse when prone than when supine.^69^ When supine, the compressive forces are a result of the gravid uterus resting on the retroperitoneal vasculature, but when prone, the compressive forces are a result of the maternal body weight compressing both the retroperitoneal vasculature and gravid uterus against the sleeping surface. While our study and studies of other investigators suggest that prone sleeping posture is absent-to-rare in the third trimester, the prevalence and effect of prone sleep in the first and second trimester remain to be elucidated. At present, sleeping posture advice in pregnancy focuses on the third trimester (i.e., supine posture should be avoided and lateral going to sleep postures should be preferred); however, additional advice regarding avoidance of prone posture as early as the second trimester may be forthcoming.

We also observed differences in the number of episodes per night, average duration of episodes, and maximum duration of episodes in various sleeping postures between the pregnant participants and bed partners. Pregnant participants had significantly more episodes and significantly longer average and maximum duration of episodes in the left lateral posture. Similarly, for right lateral posture, while they did not have a significant difference in the number of episodes per night, pregnant participants did have significantly longer average and maximum duration of episodes. They also had significantly more episodes per night and significantly longer maximum duration of episodes in the left tilt posture than their bed partners. In regard to supine posture, pregnant participants had significantly less episodes per night and significantly shorter average and maximum duration of episodes. All these results are consistent with previous reports that pregnant participants’ typical sleep has increased cumulative time spent in lateral postures and decreased cumulative time spent supine. The pregnant participants’ increased number of episodes of the lateral and left tilt postures and decreased number of episodes of the supine posture indicate an overall increased number of posture changes during sleep in late pregnancy, a preference for lateral postures, and an avoidance of the supine posture. This is likely due to conscious preferences/avoidances and a smaller number of comfortable options for sleeping posture in late pregnancy. Furthermore, in comparing our results to those from a previous video-based, in-home, third-trimester sleep study that included bed partners,^49^ the number of episodes and average duration of episodes that we observed in the left lateral, right lateral, and supine postures by the pregnant participants were remarkably similar. Finally, pregnant participants had a small but significant increase in the number of episodes of sitting per night, which we postulate may be due to various sleep disruptions in the pregnant participants that we observed while scoring their videos (e.g., sitting up to drink water, take antacids, apply lip balm, itch skin, rearrange pillows while changing postures, and apply ice to relieve carpal tunnel symptoms).

Pregnancy status (i.e., antepartum versus postpartum) impacted cumulative time spent in various sleeping postures, with postpartum participants spending more time supine (SS) and less time left lateral (SS) and right lateral (NSS) than their antepartum counterparts, and this corroborates other reports.^46,70^ More time spent sitting (SS) and longer average duration of episodes spent sitting (SS) by postpartum participants was likely due to rising to breastfeed or pump during the night.

In this study, 53.7% of pregnant participants slept on the left side of the bed. Only two studies of sleep in pregnancy report the side of the bed that the pregnant participant slept on: 43% slept on the left side of bed in Warland & Dorrian^48^ and 76.7% slept on the left side of the bed in McIntyre et al.^49^ While McIntyre et al. did not analyze the impact of the side of the bed on sleep posture or characteristics, Warland and Dorrian reported that participants who slept on the left side of the bed achieved more sleep (SS) and hypothesized this was due to more comfort sleeping on their left when facing out of the bed and away from their bed partner.^48^ They also proposed that sleeping on the left side of the bed may assist pregnant individuals in maintaining a left lateral sleeping posture although their results for this parameter were not SS. Our results do not support this assertion as we did not find an effect of the side of the bed on lateral sleeping postures in our sample of pregnant participants. The only effect we found was that right recovery posture was increased by 20.3 minutes (SS) with sleeping on the right side of the bed in late pregnancy. For bed partners, however, we confirmed Warland & Dorrian’s assertion and found that sleeping on the right side of the bed increased right lateral time by 47.9 minutes (SS) and right recovery time by 29.2 minutes (SS) and decreased left lateral time by 31.8 minutes (NSS; p 0.053), left recovery time by 10.9 minutes (NSS), and left tilt time by 13.6 minutes (SS). This result may have important clinical implications for non-pregnant adults with health conditions, such as gastroesophageal reflux disease, in which symptoms are impacted by the direction (i.e., right versus left) of lateral sleeping posture.^71^

Besides the aforementioned SS reduction in cumulative time spent in the left recovery posture, the presence of a pregnancy pillow did not impact sleeping posture of pregnant participants. On the contrary, for bed partners, the presence of a pregnancy pillow on the bed resulted in a small but SS increase in cumulative nightly sitting time (0.95 minutes). This may be due to the bed partner helping to arrange the pregnancy pillow for their pregnant partner each night. We also observed that in the presence of a pregnancy pillow, bed partners had SS increases in average and maximum duration of episodes of prone posture (13.4 minutes and 28.7 minutes, respectively). This observation may be related to the bed partner having less space on the bed in the presence of a pregnancy pillow, but we did not collect information on the size of the bed. Based on anecdotal observations that less bed sheet coverage was maintained in the presence of a pregnancy pillow, this finding may also represent the bed partner’s effort to stay warm by a combination of maximizing skin contact with the sleeping surface and breathing against the sleeping surface.

In the presence of a bed partner, in addition to less time spent in recovery postures (previously discussed), pregnant participants also spent 46.0 less minutes in right tilt (SS). Approximately 10 minutes less were spent in left tilt too (NSS). One explanation for this may be that the pregnant participant was sleeping facing away from their bed partner and either did not want to bump into their bed partner when rolling toward their back into a tilt posture or were prevented from rolling backward into a tilt posture by the bed partner’s body itself.

While sharing bed sheets did not impact the bed partners’ cumulative time in each sleeping posture, pregnant participants who shared bed sheets with their bed partner spent 94.5 more minutes in the right lateral posture (SS) and 21.2 more minutes in the left lateral posture (NSS). When the duration of episodes in each sleeping posture were considered, pregnant participants’ maximum duration of episodes in right lateral were 49.4 minutes longer (SS) with shared bed sheets, and bed partners’ maximum duration of episodes in right lateral were 21.3 minutes longer (SS) with shared bed sheets. In fact, the average and maximum durations of episodes for most postures were positive (increased) in the presence of shared bed sheets although not all were SS. One contributor to these effects may be that, in the setting of shared bed sheets, both co-sleepers feel they have less freedom to change postures and are attempting to maintain their sleeping posture for a longer duration for fear of disturbing or waking the other person when changing postures. These results suggest that shared bed sheets (as opposed to separate bed sheets or no bed sheets) may act to increase time spent sleeping in lateral postures during sleep in late pregnancy.

Pregnant participants with pets present in the sleep environment spent a cumulative of 25.4 more minutes (SS) in left tilt, whereas bed partners spent a cumulative of 50.9 more minutes (SS) in right lateral. When the duration of episodes in each sleeping posture were considered, pregnant participants’ average duration of episodes in the left lateral posture were reduced by 13 minutes (SS) in the presence of pets, whereas bed partners’ maximum duration of episodes in the right lateral posture were increased by 20.1 minutes (SS). Other than the impact of the pet occupying part of the sleeping surface and, thereby, preventing or encouraging the co-sleepers’ adoption of various sleeping postures, we cannot explain these effects.

Finally, we did not include children in the multiple linear regression models of cumulative and episodic time spent in each sleeping posture regressed on sleeping environment variables. This was because the addition of the children variable did not change the models’ R-squared, which indicates that the presence or absence of children explains 0% of the variance in the cumulative and episodic time spent in each sleeping posture. Note that only six couples (14.6%) had a child or children enter their bedroom at one or more points during one or more nights. This absence of impact on the R-squared was in contrast to the other sleeping environment variables (for example, pets) likely because the exposure to children was only transient and not continuous (for example, as in the case of pets, which slept on the bed or at the bedside). That is, when a couple was coded as having a child present, the child entered the bedroom for only a couple minutes and then left the room for the remainder of the night. We did not observe any couples where a child was continuously in the bedroom or on the bed. While the continuous presence of a child in the bedroom or on the bed was not the case for any couples in our study, if this were the case for a couple in the real world, it is likely that it would impact sleeping posture.

### Transition Postures

We observed statistically significant differences between pregnant participants and bed partners for all measures of four transition postures. With regard to the going-to-sleep posture (GTSP), pregnant participants spent more time in the GTSP prior to changing posture (45.1 versus 27.7 minutes) than the bed partners. These results corroborate those of Wilson et al.^35^ who reported a median of 51.5 minutes in the GTSP prior to changing posture and is significantly less than Stone et al.^68^ who reported a median of 62 minutes and McIntyre et al.^49^ who reported a median of 73.5 minutes. Our findings provide novel insight by inclusion of the bed partners, and given the known challenges achieving a comfortable sleeping posture, it was unexpected that the pregnant participants spent significantly more time in the GTSP prior to changing posture than the bed partners. This “postural inertia” may indicate that pregnant participants face increased difficulty changing postures, have a fewer number of comfortable postures available to them, and make more attempts to get and remain comfortable (i.e., with pillows) within the GTSP before abandoning the posture. It could also indicate that pregnant participants are more likely to achieve a deeper level of sleep initially, which enables them to maintain their GTSP for a longer duration. Pregnant participants also had more cumulative time in the GTSP (161.0 versus 125.5 minutes) and more cumulative percentage of the night in the GTSP (41.2% versus 31.1%) than the bed partners. This also corroborates Wilson et al.^35^ who reported that pregnant participants spent a cumulative 44.5% of their total sleep time (TST) in the GTSP, and it reinforces the notion that a significant proportion (but not the majority) of the night is spent in the GTSP in late pregnancy.

With regard to the waking-to-void posture (WTVP), pregnant participants spent slightly more time in the WTVP since the last posture change (29.1 versus 25.8 minutes; difference 12.4 minutes; p 0.005) than their bed partner. Pregnant participants also spent more time in the returning-to-bed posture (RTBP) prior to the next posture change (40.8 versus 19.6 minutes; difference 16.1 minutes; p 0.004). Together, these suggest that pregnant participants either have increased comfort, tolerate more discomfort, and/or make increased attempts to remain comfortable in the WTVP and RTBP before abandoning these postures. Our consideration of the WTVP and RTBP provides novel clinical insight because patients often ask about the implications of waking up at some point during the night on their back, and clinicians often just encourage them to return to sleep on their side. Clinicians can inform patients that while the median time in the WTVP is about 29 minutes, the average duration of an episode of supine sleep is only about 11.3 minutes (IQR 17.4) and the maximum duration is about 36 minutes (IQR 62.8). Furthermore, clinicians can also reassure patients that they will spend a significant amount of time (median 40.8 minutes) in the posture they settle in when they return to bed (RTBP).

With regard to the waking-in-the-morning posture (WITMP), pregnant participants spent more time in the WITMP prior to changing posture (31.2 versus 25.2 minutes), more cumulative time in the WITMP (153.4 versus 108.9 minutes), and more cumulative percentage of the night in the WITMP (38.7% versus 28.0%) than the bed partners. The latter result is slightly less but still in agreement with Wilson et al.^35^ who observed that pregnant participants spent a cumulative 45.0% of their TST in the WITMP. Furthermore, they reported that if the WITMP was supine, a median of 32.4% of the pregnant participant’s TST was spent supine, and this percentage of the night spent supine was significantly more than if the WITMP was non-supine (median 17.3%). We, however, were unable to make this delineation because our participants had multiple different WITMP’s due to completing our study over four nights, whereas Wilson et al.’s study was only a single night.

### Sleeping Posture Changes

We did not observe any SS differences in the average time in bed per night nor any sleeping posture change parameters (total number of posture changes per night, total number of posture changes per person per night, total PCI, hourly PCI’s) between pregnant participants and bed partners in the 33 couples, except for a lower PCI at hour 10 in pregnant participants. Our results, however, provide unique insights.

First, because we included bed partners, we were able to report the total number of posture change events per night for the couples (42.2 ± 15.0). On a per person basis, this represents 23.3 ± 10.3 and 22.9 ± 8.7 posture changes for the pregnant participants and the bed partners, respectively. These figures are significantly higher than any previously reported, but they mirror previously reported number of awakenings in the third trimester (15.78 ± 3.7 in Wołynczyk-Gmaj et al.^72^; 18.8 ± 6.8 in Wilson et al.^46^). For pregnant participants in the third trimester, the mean ± SD number of posture changes per night has been reported as 5.7 ± 3.9 (Lucchini et al.; excluding the first hour of recording; sensor; home-based)^67^ and 9.7 ± 4.6 (Wilson et al.; sensor in home-based participants, and video and sensor in lab-based participants)^35^ and the median number of posture changes has been reported as 6 (Kember et al.; video; lab-based),^50^ 8 (McIntyre et al.; video; home-based),^49^ and 9 (Stone et al.; video; home-based).^68^ The most probable reason that we observed a significantly higher number of posture changes per person per night is because more posture changes are detected with our thirteen-posture classification system than with the five-posture classification system used in previous studies. For example, in our study, a participant who is supine and then shifts to tilt the pelvis to the left (i.e., supine thorax with left pelvic tilt) would be deemed to have changed posture, whereas this event would not be counted as a posture change under a five-posture classification system.

Second, we looked at whether a posture change in one person was followed by no posture change or a posture change in the other person within 30 seconds, 31-60 seconds later, or 61-90 seconds later. Most posture changes by the pregnant participant and bed partner (84.5% and 84.3%, respectively) were not followed by a posture change in their co-sleeper within 90 seconds. A mean ± SD of 3.6 ± 2.0 of posture changes in one person were accompanied by a posture change in their co-sleeper within 30 seconds. In terms of percentage, about 15.5% of the pregnant participant’s posture changes were accompanied by a posture change by the bed partner within 30 seconds, whereas about 15.7% of the bed partner’s posture changes were accompanied by a posture change by the pregnant participant within 30 seconds. In the event of a posture change in one’s co-sleeper, bed partners were more likely to change their posture 31-60 seconds later or 61-90 seconds later but this did not reach SS. However, the hourly PCI correlation analysis gave further insight where simple linear regression clearly demonstrated that the bed partner’s sleeping posture was more sensitive to their co-sleeper’s posture changes than the pregnant participant’s sleeping posture (discussed later). Overall, these results suggest that the stability of a pregnant adult’s sleeping posture in response to their bed partner changing posture is equivalent or greater than that of non-pregnant adults and add to our understanding of sleep as a dyadic behaviour in co-sleeping couples.^73–77^

Third, we attempted to capture the frequency of posture changes as the night progressed by calculating the hour-by-hour PCI for the pregnant participants and bed partners and plotted them according to the absolute and normalized time elapsed (**Figure 2**). For all participants, we observed that the PCI generally followed a U-shape (**Figure 2-C**) with highest PCI occurring in the first and last hours of the overnight video recording, which reflects numerous posture changes while attempting to settle to sleep and when waking for the day, and a nadir occurring in the intervening hours. When compared with the bed partners, the pregnant participants’ mean hourly PCI was lesser at the beginning and end of the overnight video recording. While this trend was NSS, it reflects a longer duration of the GTSP and WITMP for the pregnant participants (previously discussed). Upon close inspection of the grouped mean hourly PCI against the percentage of the overnight video elapsed (**Figure 2-D**), the bed partners had two nadirs in their PCI (at 17% and 85% of the overnight video elapsed), whereas the pregnant participants had four nadirs (at 20%, 30%, 84%, and 90%). These four nadirs might also be conceptualized as two “double nadirs” each punctuated by a temporal increase in posture changes (e.g., absence from bed to void). While a trade-off resulting in less deep sleep and more light sleep in the third trimester has been well-established in the literature,^46,78–81^ these studies report cumulative amounts of light and deep sleep and do not report whether there are changes present in the distribution or timing of light and deep sleep across the night. In non-pregnant adults with healthy sleep, deep sleep is mostly concentrated in the first third-to-half of the night and decreases progressively with subsequent sleep cycles.^82^ The clustering of nadirs in the pregnant participants’ mean hourly PCI’s at the beginning and end of their overnight video recordings may reflect pregnancy-induced alterations in the timing of sleep architecture with deep sleep occurring relatively early in the first half of the night and then relatively late in the second half of the night.

We observed a statistically significant moderate positive correlation in hourly PCI’s between the pregnant participant and their bed partner. This suggests a clear relationship between co-sleepers’ posture changes: as the pregnant participant’s PCI increases, the bed partner’s PCI also increases, and vice versa. The Pearson’s R that we observed (0.37) yields an R^2^ of 0.135, which indicates that 13.5% of the variation in posture changes for an individual is explained by their co-sleeper’s posture changes. It is likely that the synchronization we observed in couples’ PCI’s was due to physical disturbance caused by co-sleeper’s movements (e.g., mattress and/or bed sheet movement as a result of posture changes) or shared environmental factors (e.g., both co-sleepers feeling too warm). Furthermore, we observed greater stability in the pregnant participant’s sleeping posture in response to posture changes in their co-sleeper and, conversely, that the bed partner’s sleeping posture was more sensitive to their co-sleeper’s posture changes. This effect is the opposite of what we expected given that sleep is known to be more light and less deep in pregnancy (previously discussed). This effect, however, may be due to the bed partner having more options for comfortable sleeping posture and, thereby, undergoing more subtle changes in posture (e.g., left lateral to supine). The bed partner may also make more efforts to create less disturbance when changing posture. In contrast, due to a restricted number of options for comfortable sleeping posture options in pregnancy, posture changes in pregnancy are often more extreme (e.g., left lateral to right lateral), require more effort and/or longer duration of time due to anatomical and biomechanical changes in pregnancy, and often require rearrangement of supporting pillows – all of which are more likely to disturb the co-sleeper.

In considering the impact of sleep environment variables on sleeping posture changes, our main finding was a drastic increase in the total number of posture changes per person per night in the presence of pets for the pregnant participants and the bed partners (17.3 and 8.0, respectively). Notably, this effect was most pronounced for the pregnant participants, which is possibly a result of pregnant participants’ sleep being lighter (previously discussed) and more likely to be disturbed when the pet moves or makes noise. This result has significant implications vis-a-vis sleep hygiene in both pregnant and non-pregnant adults and lends objective support to a mounting body of evidence implicating negative sleep effects from sharing one’s sleeping environment with a pet.^83–88^ While it might have been expected that having a bed partner and/or sharing one’s bed sheets with a bed partner would increase the total number of posture changes per person per night,^73^ our study was underpowered to detect SS effects to this regard. Supporting the so-called “Scandinavian Sleep Method”,^89^ pregnant participants who shared bed sheets with their bed partner had an average increase of 5.5 posture changes per night (p 0.09), but this effect was not observed in the opposite direction (i.e., bed partners who shared bed sheets with their pregnant partners did not have a trend of more posture changes per night). Somewhat unexpectedly, pregnant participants who slept in the presence of a bed partner had an average decrease of 7.7 posture changes per night (p 0.09). This paradoxical finding may indicate sleep-stage synchronisation (previously reported by Drews et al.)^90^ and is potentially a reflection of the known interplay between relationship quality and sleep quality in co-sleeping couples.^91–94^ Taken together, these results suggest that for pregnant adults in the third trimester, co-sleeping with one’s partner but with separate bed sheets may improve sleep by reducing the number of posture changes. Further exploration of this hypothesis is warranted.

### Sleeping Behaviour

Pregnant participants had significantly more absences from bed per night (1.3 versus 0.5) compared to their bed partners. The number of absences from bed per night for the pregnant participants in our study was similar to that reported in other studies of sleep in the third trimester.^35,49^ In late pregnancy, rising to void at least once during the night is common and accounts for the higher number of absences from bed that we observed in the pregnant participants.^95^ Furthermore, pregnant participants spent significantly more cumulative time absent from bed per night (3.2 versus 0.88 minutes) compared to their bed partners. The cumulative time spent absent from bed per night by the pregnant participants in our study is similar to a previous in-home, video-based, third-trimester study (2.5 minutes).^49^

Considering the impact of the sleep environment on sleeping behaviour, we observed that bed partners with pets present spent significantly more time (2.8 minutes) absent from bed per night than those without pets. This phenomenon was also observed in the pregnant participants but did not reach statistical significance. These results contribute further evidence regarding the negative sleep effects from sharing one’s sleeping environment with a pet (previously discussed).

### Limitations

The main limitation of our study is its relatively small sample size. Our target sample size was N=60 couples (60 pregnant participants and 60 bed partners), but only 41 pregnant participants and 36 bed partners successfully completed our study, which resulted in our study being underpowered to detect some differences had our sample size been greater. Furthermore, our participants completed the study across only four nights of the third trimester. While these nights were not required to be consecutive, most participants completed the four nights of data collection within one week. As such, our data reflects sleeping posture and behaviours over only a small segment (approximately one twelfth) of the third trimester, which may be a limitation. Sleeping posture, behaviours, and physiology are known to change significantly across the three trimesters.^8^ We are unaware of any literature, however, demonstrating that sleeping posture and behaviours change significantly within the third trimester.

While we consider sleeping posture in our study to have been prospectively (not retrospectively), continuously (all night), longitudinally (four nights), and objectively determined, scoring of body posture by video is not completely objective. For example, a lateral tilt posture with 30-60° of tilt is clearly scored as lateral tilt, whereas if the angle of tilt is more extreme than this (e.g., 15-30° or 60-75°), the posture may be better scored as supine or as a complete lateral posture. While all participants in our study wore an accelerometer on their chest, the sensor only outputs posture according to a five-posture classification, the raw signal and angle of tilt was not available to our team, and we were not privy to the angle at which the sensor classified the posture as supine versus right or left lateral. As such, visual determination of some sleeping postures (e.g., lateral tilt postures) by a trained human scorer has an inherent subjectivity, especially in circumstances in which the body is obscured (e.g., co-sleeper entanglement, thick bed sheets and blankets, low lighting).

We observed little to no time spent in twisted postures (supine thorax with left pelvic tilt, supine thorax with right pelvic tilt, supine pelvis with left thorax tilt, supine pelvis with right thorax tilt), which limited our ability to determine whether they were impacted by the sleep environment. Their low frequency of occurrence, while still higher than the frequency of prone sleeping in the pregnant participants, casts doubt on the utility of their inclusion in the thirteen-posture classification system we used. Furthermore, with regard to the sleeping environment, we did not determine the bed size (i.e., double, queen, king) or mattress type (i.e., memory foam, firm, soft, or water) and, as such, could not adjust our analyses for these variables which could have an impact on sleeping posture especially if the sleeping surface was crowded (e.g., extra support pillows, pregnancy pillow).

The sleeping posture change analysis, while accounting for posture changes in both the pregnant participant and their bed partner within the same epoch (within 30 seconds) as well as in the following two epochs (31-60 seconds and 61-90 seconds), had at least two limitations. First, it did not detect when a person moved significantly but did not ultimately change posture. Although we occasionally observed this occurring, especially following a posture change in the co-sleeping person, we did not count this as a posture change. Therefore, the true amount of sleep disruption as a result of one’s co-sleeper’s posture changes in late pregnancy may be higher than we reported. Second, regarding dyadic behaviour in co-sleeping couples, in the event of the pregnant participant and the bed partner both changing posture during the same epoch, our scoring technique did not enable us to indicate which person started changing posture first but only that both people changed posture during the same epoch (i.e., within 30 seconds of each other). As such, for these occurrences, it is not possible to know if it was the pregnant participant’s posture change that prompted the bed partner’s posture change or vice versa.

## Data Availability

All data produced in the present study are available upon reasonable request to the corresponding author.

## Acknowledgments

We thank Ectosense NV (Leuven, Belgium) for providing us with NightOwl sensors for the SLeeP AID4 Study free-of-charge. We thank all our pregnant participants and their bed partners (including those bed partners who did not participate) for their enthusiasm and dedication and for allowing us to have access to one of the most personal spaces in their homes. The data you gave us is invaluable. We thank Mr. Sina Akbarian for his mentorship role in the planning phase of this research regarding data science expertise. We thank Ms. Alexandra Gratton and Dr. Ramak Ajideh for their assistance in participant recruitment. We thank research assistants, Mr. Michael MacIntyre and Mr. Daniel MacIntyre, for their assistance in data processing.

## List of Abbreviations

ASA PS: American Society of Anesthesiologists Physical Status
GTSP: Going-to-sleep posture
IQR: Interquartile range
NSS: Not statistically significant
OSA: Obstructive sleep apnea
PCI: Posture change index
RTBP: Returning-to-bed posture
SD: Standard deviation
SLeeP AID4 Study: Sleep in Late Pregnancy: Artificial Intelligence Development for the Detection of Disturbances and Disorders Study
SS: Statistically significant
TST: Total sleep time
WITMP: Waking-in-the-morning posture
WTVP: Waking-to-void posture

## Glossary of Terms

**Absence**: An event in which a person exited the bed for the majority of an epoch (at least 15 seconds) and then subsequently returned to the bed in the same overnight video.

**Episode**: The period of time a person spends in a posture prior to changing to a different posture

**Going-to-sleep posture**: The posture that the person attempts to settle to sleep in.

**Low-risk singleton pregnancy**: An ultrasound-dated pregnancy in which one baby is carried to term with no major health issues for the mother or baby.

**Posture change index**: The average number of posture changes that a person undergoes per hour.

**Returning-to-bed posture**: The posture that the person adopts when they return to the bed following an absence from bed.

**Transition postures**: The going-to-sleep posture, waking-to-void posture, returning-to-bed posture, and waking-in-the-morning posture.

**Waking-in-the-morning posture**: The posture that the person is in immediately prior to waking for the day and exiting the bed.

**Waking-to-void posture**: The posture that the person was in immediately prior to their absence from bed.

## References

1. Mukherjee S, Patel SR, Kales SN, et al. An Official American Thoracic Society Statement: The Importance of Healthy Sleep. Recommendations and Future Priorities. Am J Respir Crit Care Med. 2015;191(12):1450–1458. doi:10.1164/rccm.201504-0767ST

2. Chaput JP, Dutil C, Featherstone R, et al. Sleep duration and health in adults: an overview of systematic reviews. Appl Physiol Nutr Metab. 2020;45(10 (Suppl. 2)):S218–S231. doi:10.1139/apnm-2020-0034

3. Li L, Zhao K, Hua J, Li S. Association between Sleep-Disordered Breathing during Pregnancy and Maternal and Fetal Outcomes: An Updated Systematic Review and Meta-Analysis. Front Neurol. 2018;9:91. doi:10.3389/fneur.2018.00091

4. Warland J, Dorrian J, Morrison JL, O’Brien LM. Maternal sleep during pregnancy and poor fetal outcomes: A scoping review of the literature with meta-analysis. Sleep Med Rev. 2018;41:197–219. doi:10.1016/j.smrv.2018.03.004

5. Lu Q, Zhang X, Wang Y, et al. Sleep disturbances during pregnancy and adverse maternal and fetal outcomes: A systematic review and meta-analysis. Sleep Med Rev. 2021;58:101436. doi:10.1016/j.smrv.2021.101436

6. Brown NT, Turner JM, Kumar S. The intrapartum and perinatal risks of sleep-disordered breathing in pregnancy: a systematic review and metaanalysis. Am J Obstet Gynecol. 2018;219(2):147–161.e1. doi:10.1016/j.ajog.2018.02.004

7. Liu L, Su G, Wang S, Zhu B. The prevalence of obstructive sleep apnea and its association with pregnancy-related health outcomes: a systematic review and meta-analysis. Sleep Breath Schlaf Atm. 2019;23(2):399–412. doi:10.1007/s11325-018-1714-7

8. Kember AJ, Elangainesan P, Ferraro ZM, Jones C, Hobson SR. Common sleep disorders in pregnancy: a review. Front Med. 2023;10. Accessed August 23, 2023. https://www.frontiersin.org/articles/10.3389/fmed.2023.1235252

9. Stacey T, Thompson JMD, Mitchell EA, Ekeroma AJ, Zuccollo JM, McCowan LME. Association between maternal sleep practices and risk of late stillbirth: a case-control study. BMJ. 2011;342:d3403. doi:10.1136/bmj.d3403

10. Owusu JT, Anderson FJ, Coleman J, et al. Association of maternal sleep practices with pre-eclampsia, low birth weight, and stillbirth among Ghanaian women. Int J Gynaecol Obstet Off Organ Int Fed Gynaecol Obstet. 2013;121(3):261–265. doi:10.1016/j.ijgo.2013.01.013

11. Gordon A, Raynes-Greenow C, Bond D, Morris J, Rawlinson W, Jeffery H. Sleep position, fetal growth restriction, and late-pregnancy stillbirth: the Sydney stillbirth study. Obstet Gynecol. 2015;125(2):347–355. doi:10.1097/AOG.0000000000000627

12. McCowan LME, Thompson JMD, Cronin RS, et al. Going to sleep in the supine position is a modifiable risk factor for late pregnancy stillbirth; Findings from the New Zealand multicentre stillbirth case-control study. PLoS ONE. 2017;12(6):e0179396. doi:10.1371/journal.pone.0179396

13. Heazell A, Li M, Budd J, et al. Association between maternal sleep practices and late stillbirth - findings from a stillbirth case-control study. BJOG Int J Obstet Gynaecol. 2018;125(2):254–262. doi:10.1111/1471-0528.14967

14. O’Brien LM, Warland J, Stacey T, Heazell AEP, Mitchell EA, STARS Consortium. Maternal sleep practices and stillbirth: Findings from an international case-control study. Birth Berkeley Calif. 2019;46(2):344–354. doi:10.1111/birt.12416

15. Cronin RS, Li M, Thompson JMD, et al. An Individual Participant Data Meta-analysis of Maternal Going-to-Sleep Position, Interactions with Fetal Vulnerability, and the Risk of Late Stillbirth. EClinicalMedicine. 2019;10:49–57. doi:10.1016/j.eclinm.2019.03.014

16. Anderson NH, Gordon A, Li M, et al. Association of Supine Going-to-Sleep Position in Late Pregnancy With Reduced Birth Weight. JAMA Netw Open. 2019;2(10):e1912614. doi:10.1001/jamanetworkopen.2019.12614

17. Lee SWY, Khaw KS, Ngan Kee WD, Leung TY, Critchley LAH. Haemodynamic effects from aortocaval compression at different angles of lateral tilt in non-labouring term pregnant women. Br J Anaesth. 2012;109(6):950–956. doi:10.1093/bja/aes349

18. Humphries A, Mirjalili SA, Tarr GP, Thompson JMD, Stone P. The effect of supine positioning on maternal hemodynamics during late pregnancy. J Matern-Fetal Neonatal Med Off J Eur Assoc Perinat Med Fed Asia Ocean Perinat Soc Int Soc Perinat Obstet. 2019;32(23):3923–3930. doi:10.1080/14767058.2018.1478958

19. Couper S, Clark A, Thompson JMD, et al. The effects of maternal position, in late gestation pregnancy, on placental blood flow and oxygenation: an MRI study. J Physiol. 2021;599(6):1901–1915. doi:10.1113/JP280569

20. Kienzl D, Berger-Kulemann V, Kasprian G, et al. Risk of inferior vena cava compression syndrome during fetal MRI in the supine position – a retrospective analysis. J Perinat Med. 2014;42(3):301–306. doi:10.1515/jpm-2013-0182

21. Higuchi H, Takagi S, Zhang K, Furui I, Ozaki M. Effect of lateral tilt angle on the volume of the abdominal aorta and inferior vena cava in pregnant and nonpregnant women determined by magnetic resonance imaging. Anesthesiology. 2015;122(2):286–293. doi:10.1097/ALN.0000000000000553

22. Stone PR, Burgess W, McIntyre JPR, et al. Effect of maternal position on fetal behavioural state and heart rate variability in healthy late gestation pregnancy. J Physiol. 2017;595(4):1213–1221. doi:10.1113/JP273201

23. Rossi A, Cornette J, Johnson MR, et al. Quantitative cardiovascular magnetic resonance in pregnant women: cross-sectional analysis of physiological parameters throughout pregnancy and the impact of the supine position. J Cardiovasc Magn Reson Off J Soc Cardiovasc Magn Reson. 2011;13:31. doi:10.1186/1532-429X-13-31

24. Khatib N, Weiner Z, Beloosesky R, Vitner D, Thaler I. The effect of maternal supine position on umbilical and cerebral blood flow indices. Eur J Obstet Gynecol Reprod Biol. 2014;175:112–114. doi:10.1016/j.ejogrb.2013.12.043

25. Robertson N, Okano S, Kumar S. Sleep in the Supine Position During Pregnancy is Associated with Fetal Cerebral Redistribution. J Clin Med. 2020;9(6):1773. doi:10.3390/jcm9061773

26. Nakai Y, Mine M, Nishio J, Maeda T, Imanaka M, Ogita S. Effects of maternal prone position on the umbilical arterial flow. Acta Obstet Gynecol Scand. 1998;77(10):967–969.

27. van Katwijk C, Wladimiroff JW. Effect of maternal posture on the umbilical artery flow velocity waveform. Ultrasound Med Biol. 1991;17(7):683–685. doi:10.1016/0301-5629(91)90099-i

28. Zun Z, Zaharchuk G, Andescavage NN, Donofrio MT, Limperopoulos C. Non-Invasive Placental Perfusion Imaging in Pregnancies Complicated by Fetal Heart Disease Using Velocity-Selective Arterial Spin Labeled MRI. Sci Rep. 2017;7(1):16126. doi:10.1038/s41598-017-16461-8

29. Silva KP, Hamamoto TENK, Nomura RMY. Transient fetal blood redistribution associated with maternal supine position. J Perinat Med. 2017;45(3):343–347. doi:10.1515/jpm-2016-0288

30. Abaci Turk E, Abulnaga SM, Luo J, et al. Placental MRI: Effect of maternal position and uterine contractions on placental BOLD MRI measurements. Placenta. 2020;95:69–77. doi:10.1016/j.placenta.2020.04.008

31. Aldrich CJ, D’Antona D, Spencer JA, et al. The effect of maternal posture on fetal cerebral oxygenation during labour. Br J Obstet Gynaecol. 1995;102(1):14–19. doi:10.1111/j.1471-0528.1995.tb09019.x

32. Oksenberg A, Gadoth N. Are we missing a simple treatment for most adult sleep apnea patients? The avoidance of the supine sleep position. J Sleep Res. 2014;23(2):204–210. doi:10.1111/jsr.12097

33. Loube DI, Poceta JS, Morales MC, Peacock MD, Mitler MM. Self-reported snoring in pregnancy. Association with fetal outcome. Chest. 1996;109(4):885–889. doi:10.1378/chest.109.4.885

34. Warland J, Dorrian J, Kember AJ, et al. Modifying Maternal Sleep Position in Late Pregnancy Through Positional Therapy: A Feasibility Study. J Clin Sleep Med JCSM Off Publ Am Acad Sleep Med. 2018;14(8):1387–1397. doi:10.5664/jcsm.7280

35. Wilson DL, Fung AM, Pell G, et al. Polysomnographic analysis of maternal sleep position and its relationship to pregnancy complications and sleep-disordered breathing. Sleep. 2022;45(4):zsac032. doi:10.1093/sleep/zsac032

36. Wilson DL, Whenn C, Barnes M, Walker SP, Howard ME. A Position Modification Device for the Prevention of Supine Sleep During Pregnancy: A Randomised Crossover Trial. BJOG Int J Obstet Gynaecol. Published online September 16, 2024. doi:10.1111/1471-0528.17952

37. Kember AJ, Anderson JL, Gorazd NE, et al. Maternal posture-physiology interactions in human pregnancy: a narrative review. Front Physiol. 2024;15. doi:10.3389/fphys.2024.1370079

38. Kember AJ, Anderson JL, House SC, Reuter DG, Goergen CJ, Hobson SR. Impact of maternal posture on fetal physiology in human pregnancy: a narrative review. Front Physiol. 2024;15. doi:10.3389/fphys.2024.1394707

39. Archer TL. Cardiac output-guided maternal positioning may protect the fetal oxygen supply and thereby reduce pregnancy complications. J Perinat Med. Published online October 1, 2025. doi:10.1515/jpm-2025-0333

40. Herring SJ, Foster GD, Pien GW, et al. Do pregnant women accurately report sleep time? Sleep Breath Schlaf Atm. 2013;17(4):10.1007/s11325-013-0835-2. doi:10.1007/s11325-013-0835-2

41. Wilson DL, Fung A, Walker SP, Barnes M. Subjective reports versus objective measurement of sleep latency and sleep duration in pregnancy. Behav Sleep Med. 2013;11(3):207–221. doi:10.1080/15402002.2012.670674

42. Russo K, Bianchi MT. How Reliable Is Self-Reported Body Position during Sleep? J Clin Sleep Med JCSM Off Publ Am Acad Sleep Med. 2016;12(1):127–128. doi:10.5664/jcsm.5410

43. Kai-Ching Yu C. Why is Self-Report of Sleep Position Sometimes Unreliable? Sleep Hypn - Int J. Published online May 22, 2017. doi:10.5350/Sleep.Hypn.2017.19.0140

44. Skarpsno ES, Mork PJ, Nilsen TIL, Holtermann A. Sleep positions and nocturnal body movements based on free-living accelerometer recordings: association with demographics, lifestyle, and insomnia symptoms. Nat Sci Sleep. 2017;9:267–275. doi:10.2147/NSS.S145777

45. Maasilta P, Bachour A, Teramo K, Polo O, Laitinen LA. Sleep-related disordered breathing during pregnancy in obese women. Chest. 2001;120(5):1448–1454. doi:10.1378/chest.120.5.1448

46. Wilson DL, Barnes M, Ellett L, Permezel M, Jackson M, Crowe SF. Decreased sleep efficiency, increased wake after sleep onset and increased cortical arousals in late pregnancy. Aust N Z J Obstet Gynaecol. 2011;51(1):38–46. doi:10.1111/j.1479-828X.2010.01252.x

47. O’Brien LM, Warland J. Typical sleep positions in pregnant women. Early Hum Dev. 2014;90(6):315–317. doi:10.1016/j.earlhumdev.2014.03.001

48. Warland J, Dorrian J. Accuracy of Self-Reported Sleep Position in Late Pregnancy. PLOS ONE. 2014;9(12):e115760. doi:10.1371/journal.pone.0115760

49. McIntyre JPR, Ingham CM, Hutchinson BL, et al. A description of sleep behaviour in healthy late pregnancy, and the accuracy of self-reports. BMC Pregnancy Childbirth. 2016;16(1):115. doi:10.1186/s12884-016-0905-0

50. Kember AJ, Scott HM, O’Brien LM, et al. Modifying maternal sleep position in the third trimester of pregnancy with positional therapy: a randomised pilot trial. BMJ Open. 2018;8(8):e020256. doi:10.1136/bmjopen-2017-020256

51. Dunietz GL, Sever O, DeRowe A, Tauman R. Sleep position and breathing in late pregnancy and perinatal outcomes. J Clin Sleep Med. 2020;16(6):955–959. doi:10.5664/jcsm.8416

52. Kember AJ, Zia H, Elangainesan P, et al. Transitioning sleeping position detection in late pregnancy using computer vision from controlled to real-world settings: an observational study. Published online August 23, 2023. doi:10.22541/au.169279284.45458103/v1

53. Statement on ASA Physical Status Classification System. American Society of Anesthesiologists. December 13, 2020. Accessed December 10, 2025. https://www.asahq.org/standards-and-practice-parameters/statement-on-asa-physical-status-classification-system

54. Kember AJ, Selvarajan R, Park E, et al. Vision-based detection and quantification of maternal sleeping position in the third trimester of pregnancy in the home setting–Building the dataset and model. PLOS Digit Health. 2023;2(10):e0000353. doi:10.1371/journal.pdig.0000353

55. Peduzzi P, Concato J, Kemper E, Holford TR, Feinstein AR. A simulation study of the number of events per variable in logistic regression analysis. J Clin Epidemiol. 1996;49(12):1373–1379. doi:10.1016/s0895-4356(96)00236-3

56. Concato J, Peduzzi P, Holford TR, Feinstein AR. Importance of events per independent variable in proportional hazards analysis. I. Background, goals, and general strategy. J Clin Epidemiol. 1995;48(12):1495–1501. doi:10.1016/0895-4356(95)00510-2

57. Kember A, Ritchie L, Zia H, et al. Sleep physiology in late pregnancy: A video-based, multi-night, in-home, level 3 sleep apnea study of pregnant participants and their bed partners.

58. Fast A, Weiss L, Parikh S, Hertz G. Night backache in pregnancy. Hypothetical pathophysiological mechanisms. Am J Phys Med Rehabil. 1989;68(5):227–229. doi:10.1097/00002060-198910000-00005

59. Fast A, Shapiro D, Ducommun EJ, Friedmann LW, Bouklas T, Floman Y. Low-back pain in pregnancy. Spine. 1987;12(4):368–371. doi:10.1097/00007632-198705000-00011

60. Fast A, Hertz G. Nocturnal low back pain in pregnancy: polysomnographic correlates. Am J Reprod Immunol N Y N 1989. 1992;28(3-4):251–253. doi:10.1111/j.1600-0897.1992.tb00807.x

61. Paksoy Y, Gormus N. Epidural venous plexus enlargements presenting with radiculopathy and back pain in patients with inferior vena cava obstruction or occlusion. Spine. 2004;29(21):2419–2424. doi:10.1097/01.brs.0000144354.36449.2f

62. Sleep On Side - a pregnancy campaign Tommy’s. Accessed November 28, 2025. https://www.tommys.org/pregnancy-information/im-pregnant/sleep-side/sleep-side-pregnancy-campaign

63. Andrews CJ, Ellwood DA, Gordon A, et al. Stillbirth in Australia 2: Working together to reduce stillbirth in Australia: The Safer Baby Bundle initiative. Women Birth J Aust Coll Midwives. 2020;33(6):514–519. doi:10.1016/j.wombi.2020.09.006

64. Sleep On Side. Accessed January 11, 2023. https://www.sleeponside.org.nz/

65. National Institute for Health and Care Excellence, National Guideline Alliance, Royal College of Obstetricians and Gynaecologists. Antenatal Care: [W] Maternal Sleep Position during Pregnancy - NICE Guideline NG201 - Evidence Reviews Underpinning Recommendations 1.3.24 to 1.3.25.; 2021. Accessed May 24, 2022. https://www.ncbi.nlm.nih.gov/books/NBK573947/pdf/Bookshelf_NBK573947.pdf

66. Kember AJ, Gilad N, Hobson SR. Sleeping posture in pregnancy. CMAJ. 2025;197(23):E645–E645. doi:10.1503/cmaj.241858

67. Lucchini M, Wapner RJ, Chia-Ling NC, et al. Effects of maternal sleep position on fetal and maternal heart rate patterns using overnight home fetal ECG recordings. Int J Gynaecol Obstet Off Organ Int Fed Gynaecol Obstet. 2020;149(1):82–87. doi:10.1002/ijgo.13096

68. Stone PR, Burgess W, McIntyre J, et al. An investigation of fetal behavioural states during maternal sleep in healthy late gestation pregnancy: an observational study. J Physiol. 2017;595(24):7441–7450. doi:10.1113/JP275084

69. Ormesher L, Catchpole J, Peacock L, et al. The effect of prone positioning on maternal haemodynamics and fetal wellbeing in the third trimester–A primary cohort study with a scoping review. PLOS ONE. 2023;18(10):e0287804. doi:10.1371/journal.pone.0287804

70. Mills GH, Chaffe AG. Sleeping positions adopted by pregnant women of more than 30 weeks gestation. Anaesthesia. 1994;49(3):249–250. doi:10.1111/j.1365-2044.1994.tb03433.x

71. Simadibrata DM, Lesmana E, Amangku BR, Wardoyo MP, Simadibrata M. Left lateral decubitus sleeping position is associated with improved gastroesophageal reflux disease symptoms: A systematic review and meta-analysis. World J Clin Cases. 2023;11(30):7329–7336. doi:10.12998/wjcc.v11.i30.7329

72. Wołyńczyk-Gmaj D, Majewska A, Bramorska A, et al. Cognitive Function Decline in the Third Trimester of Pregnancy Is Associated with Sleep Fragmentation. J Clin Med. 2022;11(19):5607. doi:10.3390/jcm11195607

73. Pankhurst FP, Horne JA. The influence of bed partners on movement during sleep. Sleep. 1994;17(4):308–315. doi:10.1093/sleep/17.4.308

74. Walters EM, Phillips AJK, Boardman JM, Norton PJ, Drummond SPA. Vulnerability and resistance to sleep disruption by a partner: A study of bed-sharing couples. Sleep Health. 2020;6(4):506–512. doi:10.1016/j.sleh.2019.12.005

75. Walters EM, Phillips AJK, Mellor A, et al. Sleep and wake are shared and transmitted between individuals with insomnia and their bed-sharing partners. Sleep. 2020;43(1):zsz206. doi:10.1093/sleep/zsz206

76. Walters EM, Phillips AJ, Hamill K, Norton PJ, Drummond SP. Anxiety predicts dyadic sleep characteristics in couples experiencing insomnia but not in couples without sleep disorders. J Affect Disord. 2020;273:122–130. doi:10.1016/j.jad.2020.04.031

77. Gunn HE, Lee S, Eberhardt KR, Buxton OM, Troxel WM. Nightly sleep-wake concordance and daily marital interactions. Sleep Health. 2021;7(2):266–272. doi:10.1016/j.sleh.2020.11.003

78. Hertz G, Fast A, Feinsilver SH, Albertario CL, Schulman H, Fein AM. Sleep in normal late pregnancy. Sleep. 1992;15(3):246–251. doi:10.1093/sleep/15.3.246

79. Izci-Balserak B, Keenan BT, Corbitt C, Staley B, Perlis M, Pien GW. Changes in Sleep Characteristics and Breathing Parameters During Sleep in Early and Late Pregnancy. J Clin Sleep Med JCSM Off Publ Am Acad Sleep Med. 2018;14(7):1161–1168. doi:10.5664/jcsm.7216

80. Lee KA, Zaffke ME, McEnany G. Parity and sleep patterns during and after pregnancy. Obstet Gynecol. 2000;95(1):14–18. doi:10.1016/s0029-7844(99)00486-x

81. Schorr SJ, Chawla A, Devidas M, Sullivan CA, Naef RW, Morrison JC. Sleep patterns in pregnancy: a longitudinal study of polysomnography recordings during pregnancy. J Perinatol Off J Calif Perinat Assoc. 1998;18(6 Pt 1):427–430.

82. Bes F, Schulz H, Navelet Y, Salzarulo P. The distribution of slow-wave sleep across the night: a comparison for infants, children, and adults. Sleep. 1991;14(1):5–12. doi:10.1093/sleep/14.1.5

83. Patel SI, Miller BW, Kosiorek HE, Parish JM, Lyng PJ, Krahn LE. The Effect of Dogs on Human Sleep in the Home Sleep Environment. Mayo Clin Proc. 2017;92(9):1368–1372. doi:10.1016/j.mayocp.2017.06.014

84. Smith BP, Hazelton PC, Thompson KR, Trigg JL, Etherton HC, Blunden SL. A Multispecies Approach to Co-Sleeping : Integrating Human-Animal Co-Sleeping Practices into Our Understanding of Human Sleep. Hum Nat Hawthorne N. 2017;28(3):255–273. doi:10.1007/s12110-017-9290-2

85. Hoffman CL, Browne M, Smith BP. Human-Animal Co-Sleeping: An Actigraphy-Based Assessment of Dogs’ Impacts on Women’s Nighttime Movements. Anim Open Access J MDPI. 2020;10(2):278. doi:10.3390/ani10020278

86. Hoffman CL, Stutz K, Vasilopoulos T. An Examination of Adult Women’s Sleep Quality and Sleep Routines in Relation to Pet Ownership and Bedsharing. Anthrozoös. 2018;31(6):711–725. doi:10.1080/08927936.2018.1529354

87. Chin BN, Singh T, Carothers AS. Co-sleeping with pets, stress, and sleep in a nationally-representative sample of United States adults. Sci Rep. 2024;14(1):5577. doi:10.1038/s41598-024-56055-9

88. Bolstad CJ, Nadorff MR. Dog Tired: A Cross-Sectional Examination of the Relation Between Dog and/or Cat Ownership and Owners’ Sleep. J Sleep Res. Published online September 8, 2025:e70188. doi:10.1111/jsr.70188

89. What Is the Scandinavian Sleep Method? Sleep Foundation. June 8, 2023. Accessed November 29, 2025. https://www.sleepfoundation.org/sleep-hygiene/scandinavian-sleep-method

90. Drews HJ, Wallot S, Brysch P, et al. Bed-Sharing in Couples Is Associated With Increased and Stabilized REM Sleep and Sleep-Stage Synchronization. Front Psychiatry. 2020;11:583. doi:10.3389/fpsyt.2020.00583

91. Andersen ML, Alvarenga TA, Fonseca LH, Tufik S. Interactions between co-sleeping, sleep quality, and relationship health: The benefits and challenges of shared sleep environments. J Health Psychol. Published online August 7, 2025:13591053251356355. doi:10.1177/13591053251356355

92. Troxel WM, Robles TF, Hall M, Buysse DJ. Marital quality and the marital bed: Examining the covariation between relationship quality and sleep. Sleep Med Rev. 2007;11(5):389–404. doi:10.1016/j.smrv.2007.05.002

93. Richter K, Adam S, Geiss L, Peter L, Niklewski G. Two in a bed: The influence of couple sleeping and chronotypes on relationship and sleep. An overview. Chronobiol Int. 2016;33(10):1464–1472. doi:10.1080/07420528.2016.1220388

94. Wang XX, Lin Q, Liu X, et al. The association between couple relationships and sleep: A systematic review and meta-analysis. Sleep Med Rev. 2025;79:102018. doi:10.1016/j.smrv.2024.102018

95. Khosla L, Huang AJ, Kasarla N, Monaghan TF, Weiss JP, Kabarriti AE. Association between pregnancy and nocturia: A National Health and Nutrition Examination Survey analysis. Neurourol Urodyn. 2022;41(6):1505–1510. doi:10.1002/nau.24998

96. Elsey T, Keller PS, El-Sheikh M. The role of couple sleep concordance in sleep quality: Attachment as a moderator of associations. J Sleep Res. 2019;28(5):e12825. doi:10.1111/jsr.12825

97. Doerr JM, Klaus K, Troxel W, et al. The Effect of Intranasal Oxytocin on the Association Between Couple Interaction and Sleep: A Placebo-Controlled Study. Psychosom Med. 2022;84(6):727–737. doi:10.1097/PSY.0000000000001091

98. Troxel WM, Buysse DJ, Matthews KA, et al. Marital/cohabitation status and history in relation to sleep in midlife women. Sleep. 2010;33(7):973–981. doi:10.1093/sleep/33.7.973

99. Drews HJ, Drews A. Couple Relationships Are Associated With Increased REM Sleep-A Proof-of-Concept Analysis of a Large Dataset Using Ambulatory Polysomnography. Front Psychiatry. 2021;12:641102. doi:10.3389/fpsyt.2021.641102

