## Supplementary File for "Sleeping posture, behaviour, and environment in late pregnancy: A comprehensive analysis of a video-based, multi-night, in-home, level 3 sleep apnea study of pregnant participants and their bed partners"

#### List of Captions for Supplementary Tables

**Table S1**. Cumulative number of minutes per night and cumulative percentage of the night spent in each posture for the postpartum participants and bed partners based on video analysis

**Table S2**. Average number of episodes in each posture per night, average length (in minutes) of episodes spent in each posture, and average maximum length (in minutes) of episodes spent in each posture for the postpartum participants and bed partners based on video analysis

**Table S3**. Immediate and cumulative nightly amount of time spent in transition postures for the postpartum participants and bed partners based on video analysis

**Table S4**. Average time in bed per night, number of posture change events per night, number of posture changes per person per night, posture change interactions, and hour-by-hour posture change index for the postpartum participants and bed partners based on video analysis

**Table S5**. Cumulative number of absences per night and cumulative time (in minutes) spent absent from bed per night for the postpartum participants and bed partners based on video

**Table S6**. Cumulative number of minutes per night and cumulative percentage of the night spent in each posture for the pregnant participant and bed partner based on video analysis and NightOwl data

**Table S7**. Average time in bed per night, number of posture changes per person per night, total posture change index, and hour-by-hour posture change index for the pregnant participants sleeping with and without their bed partner based on video analysis

#### Postpartum Data Analysis

##### Cumulative Time Analysis

The cumulative number of minutes per night and cumulative percentage of the night spent in each posture by the postpartum participants and bed partners based on analysis of the overnight video recordings is shown in **Table S1**. There were no statistically significant differences between the postpartum participants and bed partners in any of these measures (difference testing not shown).

Table S1 | **Cumulative number of minutes per night and cumulative percentage of the night spent in each posture for the postpartum participants and bed partners based on video analysis**

| | **Postpartum** (n=4) | **Bed partner** (n=4) |
|---|---|---|
| *Left recovery* | | |
| Minutes | 2.2 (10.7) | 0.0 (10.5) |
| Percent of night | 2.2 ± 3.3 | 0.0 (3.9) |
| *Left lateral* | | |
| Minutes | 75.6 ± 38.9 | 111.5 ± 29.6 |
| Percent of night | 21.1 ± 15.3 | 31.1 ± 13.7 |
| *Left tilt* | | |
| Minutes | 0.08 (0.88) | 1.2 (10.7) |
| Percent of night | 0.02 (0.19) | 0.39 (3.8) |
| *Supine* | | |
| Minutes | 176.8 ± 117.4 | 132.8 ± 83.3 |
| Percent of night | 44.0 ± 27.3 | 31.5 ± 14.9 |
| *Supine thorax, right pelvis* | | |
| Minutes | 3.9 ± 5.2 | 0.08 (5.4) |
| Percent of night | 1.2 ± 1.8 | 0.03 (1.2) |
| *Supine thorax, left pelvis* | | |
| Minutes | 14.9 ± 20.1 | 0.33 (2.3) |
| Percent of night | 3.2 ± 4.4 | 0.13 (0.57) |
| *Supine pelvis, right thorax* | | |
| Minutes | 0 (0) | 0 (0) |
| Percent of night | 0 (0) | 0 (0) |
| *Supine pelvis, left thorax* | | |
| Minutes | 0 (0) | 0 (0) |
| Percent of night | 0 (0) | 0 (0) |
| *Right tilt* | | |
| Minutes | 0.0 (4.0) | 17.0 ± 23.7 |
| Percent of night | 0.0 (1.3) | 1.7 (5.1) |
| *Right lateral* | | |
| Minutes | 77.2 ± 60.4 | 88.8 ± 43.0 |
| Percent of night | 20.5 ± 17.3 | 22.7 ± 11.1 |
| *Right recovery* | | |
| Minutes | 10.9 ± 9.9 | 0.0 (3.9) |
| Percent of night | 3.0 ± 2.9 | 0.0 (1.0) |
| *Prone* | | |
| Minutes | 0 (0) | 0 (0) |
| Percent of night | 0 (0) | 0 (0) |
| *Sitting* | | |
| Minutes | 0.33 (16.1) | 0.0 (0.04) |
| Percent of night | 0.09 (3.5) | 0.0 (0.01) |

Normally distributed variables are presented as mean ± standard deviation. Non-normally distributed variables are presented as median (interquartile range).

#### Episodic Time Analysis

For each posture, the average number of episodes spent in that posture per night, the average length of these episodes, and the average maximum length of these episodes, is given in **Table S2** for the postpartum participants and bed partners based on analysis of the overnight videos. There were no statistically significant differences between the postpartum participants and bed partners in any of these measures (difference testing not shown).

Table S2 | **Average number of episodes in each posture per night, average length (in minutes) of episodes spent in each posture, and average maximum length (in minutes) of episodes spent in each posture for the postpartum participants and bed partners based on video analysis**

| | **Postpartum** (n=4) | **Bed partner** (n=4) |
|---|---|---|
| *Left recovery episodes* | [1] | [3] |
| Avg. number per night | 0.67 ± 0.33 | 1.7 (0.0) |
| Avg. length (minutes) | 15.5 ± 17.3 | 25.3 (0.0) |
| Avg. max. length (minutes) | 25.5 ± 33.6 | 61.5 (0.0) |
| *Left lateral episodes* | | |
| Avg. number per night | 3.6 ± 2.2 | 3.4 ± 1.4 |
| Avg. length (minutes) | 23.6 ± 11.0 | 34.9 ± 8.6 |
| Avg. max. length (minutes) | 76.4 ± 24.7 | 78.9 ± 14.9 |
| *Left tilt episodes* | [2] | [1] |
| Avg. number per night | 0.33 ± 0.00 | 0.33 (1.8) |
| Avg. length (minutes) | 4.8 ± 6.0 | 5.5 ± 4.2 |
| Avg. max. length (minutes) | 4.8 ± 6.0 | 10.7 ± 12.7 |
| *Supine episodes* | | |
| Avg. number per night | 5.6 ± 2.5 | 4.6 ± 1.7 |
| Avg. length (minutes) | 29.2 ± 15.5 | 31.1 ± 21.1 |
| Avg. max. length (minutes) | 113.3 ± 73.1 | 95.5 ± 49.5 |
| *STRP episodes* | [2] | [2] |
| Avg. number per night | 0.50 ± 0.24 | 1.2 ± 1.2 |
| Avg. length (minutes) | 15.0 ± 2.1 | 5.5 ± 7.1 |
| Avg. max. length (minutes) | 21.3 ± 11.0 | 24.8 ± 34.3 |
| *STLP episodes* | [2] | [2] |
| Avg. number per night | 1.7 ± 0.94 | 0.50 ± 0.24 |
| Avg. length (minutes) | 24.9 ± 24.9 | 11.0 ± 14.1 |
| Avg. max. length (minutes) | 40.8 ± 31.5 | 11.0 ± 14.1 |
| *SPRT episodes* | [4] | [4] |
| Avg. number per night | n/a | n/a |
| Avg. length (minutes) | n/a | n/a |
| Avg. max. length (minutes) | n/a | n/a |
| *SPLT episodes* | [4] | [4] |

| | | |
|---|---|---|
| Avg. number per night | n/a | n/a |
| Avg. length (minutes) | n/a | n/a |
| Avg. max. length (minutes) | n/a | n/a |
| *Right tilt episodes* | [3] | |
| Avg. number per night | 2.3 (0.0) | 1.0 ± 0.82 |
| Avg. length (minutes) | 6.8 (0.0) | 11.8 ± 10.2 |
| Avg. max. length (minutes) | 18.0 (0.0) | 23.4 ± 23.9 |
| *Right lateral episodes* | | |
| Avg. number per night | 2.8 ± 1.6 | 3.3 ± 1.4 |
| Avg. length (minutes) | 30.2 (10.4) | 26.5 ± 4.4 |
| Avg. max. length (minutes) | 69.3 ± 51.8 | 58.0 ± 11.0 |
| *Right recovery episodes* | [1] | [3] |
| Avg. number per night | 0.67 ± 0.33 | 1.0 (0.0) |
| Avg. length (minutes) | 21.3 ± 8.0 | 15.7 (0.0) |
| Avg. max. length (minutes) | 30.3 ± 17.4 | 32.5 (0.0) |
| *Prone episodes* | [4] | [4] |
| Avg. number per night | n/a | n/a |
| Avg. length (minutes) | n/a | n/a |
| Avg. max. length (minutes) | n/a | n/a |
| *Sitting episodes* | [2] | [3] |
| Avg. number per night | 2.2 ± 1.7 | 0.33 (0.00) |
| Avg. length (minutes) | 9.7 ± 12.8 | 0.50 (0.00) |
| Avg. max. length (minutes) | 20.8 ± 27.9 | 0.50 (0.00) |

Normally distributed variables are presented as mean ± standard deviation. Non-normally distributed variables are presented as median (interquartile range). Square brackets with a number enclosed, [number], indicates the number of missing values (i.e., the number of participants that did not have any episodes in the posture).
**Abbreviations**: Avg. indicates average; Max. indicates maximum; STRP indicates supine thorax, right pelvis; STLP indicates supine thorax, left pelvis; SPRT indicates supine pelvis, right thorax; SPLT indicates supine pelvis, left thorax.

#### Transition Postures Analysis

For each postpartum participant and bed partner, transition postures were analyzed to determine the length of time spent in the transition posture prior to changing to another posture (going-to-sleep posture, returning to bed posture) or since assuming that posture (waking-to-void posture, waking-in-the-morning posture) (see **Table S3**). Furthermore, for the going-to-sleep and waking-in-the-morning postures, the cumulative number of minutes per night and corresponding percentage of the night spent in each posture was determined. There were no statistically significant differences between the postpartum participants and bed partners in any of these measures (difference testing not shown).

**Table S3 | Immediate and cumulative nightly amount of time spent in transition postures for the postpartum participants and bed partners based on video analysis**

| | **Postpartum** (n=4) | **Bed partner** (n=4) |
|---|---|---|
| *Going-to-sleep posture* | | |
| Minutes in GTSP before change | 99.0 ± 45.6 | 40.7 ± 22.2 |
| Cumulative time in GTSP (minutes) | 173.1 ± 92.2 | 137.9 ± 84.0 |
| Cumulative percent of night in GTSP | 44.1 ± 22.4 | 32.5 ± 15.9 |
| *Waking-to-void posture* | [1] | [2] |
| Minutes in WTVP since last change | 15.4 ± 9.2 | 18.3 ± 19.8 |
| *Returning-to-bed posture* | [1] | [2] |
| Minutes in RTBP before change | 31.5 ± 34.5 | 27.4 ± 9.1 |
| *Waking-in-the-morning posture* | | |
| Minutes in WITMP since last change | 21.9 ± 3.9 | 31.8 ± 10.5 |
| Cumulative time in WITMP (minutes) | 116.7 ± 29.2 | 108.6 ± 35.0 |
| Cumulative percent of night in WITMP | 29.4 ± 8.9 | 31.5 ± 13.9 |

Normally distributed variables are presented as mean ± standard deviation. Non-normally distributed variables are presented as median (interquartile range). Square brackets with a number enclosed, [number], indicates the number of missing values.
**Abbreviations**: GTSP indicates going-to-sleep posture; WTVP indicates waking-to-void posture; RTBP indicates returning-to-bed posture; WITMP indicates waking-in-the-morning posture.

#### Sleeping Posture Changes

Overnight videos were analyzed to determine, for the four postpartum participant and bed partner couples, the average time in bed per night, the total number of posture change events (i.e., in which one or both co-sleepers changed posture) per night and the total number of posture changes per person per night (see **Table S4**). For each posture change event in the four couples, we indicated whether the co-sleeper did not change posture, changed posture within 30 seconds, 31-60 seconds, or 61-90 seconds. For the postpartum participant and bed partner, we provided the total nightly posture change index (PCI; the number of posture changes occurring per hour) and hour-by-hour PCI from the first hour through the last hour of the overnight video recordings to show how the frequency of posture changes varies throughout the night. There were no statistically significant differences between the postpartum participants and bed partners in any of these measures (difference testing not shown). Furthermore, we compared these measures between pregnant and postpartum participants and their bed partners and were underpowered to demonstrate any significant difference except for the PCI for hour 3 for the postpartum participants, which was significantly less than the PCI for hour 3 for the pregnant participants (difference -2.3; 95% CI -4.2 to -0.37; p 0.02; refer to **Table 5** in the main manuscript for the pregnant participant values). That said, there was a trend of lower total number of posture change events per night, lower total number of posture changes per person per night, and lower total PCI in the postpartum couples compared to the pregnant couples, possibly reflecting improved comfort and deeper sleep in the postpartum period compared to the antepartum period.

Table S4 | **Average time in bed per night, number of posture change events per night, number of posture changes per person per night, posture change interactions, and hour-by-hour posture change index for the postpartum participants and bed partners based on video analysis**

| | **Postpartum** (n=4) | **Bed partner** (n=4) |
|---|---|---|
| Average time in bed per night (hours) | 7.4 ± 1.6 | 7.2 ± 1.4 |
| Total number of posture change events per night[a] | 29.4 ± 4.9 | |
| Total number of posture changes per person per night[b] | 16.0 ± 3.8 | 15.3 ± 2.8 |
| *Number of posture changes per person per night in which the co-sleeper:* | | |
| Did not change posture | 14.1 ± 4.1 | 13.4 ± 3.2 |
| Changed posture within 30s[c] | 1.9 ± 0.50 | |
| Changed posture 31-60s later | 1.0 ± 0.47 | 0.58 ± 0.50 |
| Changed posture 61-90s later | 0.33 ± 0.27 | 0.33 (0.08) |
| *Posture change index* | | |
| Total | 2.5 ± 0.44 | 2.5 ± 0.73 |
| Hour 1 | 3.8 ± 3.3 | 1.4 (3.4) |
| Hour 2 | 1.3 ± 1.2 | 2.3 ± 0.91 |
| Hour 3 | 1.2 ± 0.92 | 2.8 ± 1.3 |
| Hour 4 | 1.9 ± 1.3 | 1.7 ± 1.4 |
| Hour 5 | 3.5 ± 2.4 | 3.0 ± 2.8 |
| Hour 6 | 2.8 (0.69) | 2.7 ± 1.8 |
| Hour 7 | 2.3 ± 1.3 | 3.0 ± 1.5 |
| Hour 8 | 2.0 ± 1.2 | 3.5 ± 1.9 |
| Hour 9 | 3.2 ± 2.4 [1] | 6.5 ± 6.3 [2] |
| Hour 10 | 5.6 (0.0) [3] | 4.9 (0.0) [3] |

Normally distributed variables are presented as mean ± standard deviation. Non-normally distributed variables are presented as median (interquartile range). Square brackets with a number enclosed, [number], indicates the number of missing values.
[a]Includes events in which the pregnant participant changed posture, bed partner changed posture, or both co-sleepers changed posture at the same time (within 30 seconds of each other).
[b]Includes all events in which the person changed posture (whether they changed posture alone or at the same time [within 30 seconds] as their co-sleeper).
[c]Our video scoring technique did not enable us to determine which co-sleeper started changing

posture first but only that both co-sleepers changed posture during the same epoch, which is why this parameter is the same for the pregnant participant and the bed partner.

#### Sleep Behaviour Characteristics

The cumulative number of absences per night and cumulative time absent from bed per night for the postpartum participants and bed partners based on analysis of the overnight video recordings is shown in **Table S5**. There were no statistically significant differences between the postpartum participants and bed partners in any of these measures (difference testing not shown).

Table S5 | **Cumulative number of absences per night and cumulative time (in minutes) spent absent from bed per night for the postpartum participants and bed partners based on video**

| | **Postpartum** (n=4) | **Bed partner** (n=4) |
|---|---|---|
| Cumulative number of absences | 0.50 ± 0.58 | 0.50 ± 0.64 |
| Cumulative time (minutes) absent | 0.42 (2.2) | 1.1 ± 1.6 |

Normally distributed variables are presented as mean ± standard deviation. Non-normally distributed variables are presented as median (interquartile range).

### NightOwl-Based Sleeping Posture Data

We compared the NightOwl-based sleeping posture data to the video-based sleeping posture analysis (see **Table S6**). We were able to make comparisons between the two approaches in the cumulative amount of minutes per night and the cumulative percentage of the night for the pregnant participants and bed partners in four postures that the NightOwl measured (left lateral, supine, right lateral, and prone). Participants (and their bed partner, if applicable) who completed the sleep studies postpartum were excluded from this analysis. Also note that two bed partners participated in the video recording aspect of the study but did not don the NightOwl sensors, so the n=31 bed partners for the NightOwl posture data instead of n=33.

For the pregnant participants, compared to video-based posture, the NightOwl significantly underestimated the cumulative number of minutes per night in the left lateral and right lateral postures by 18.9 and 27.1 minutes, respectively. Furthermore, the NightOwl significantly overestimated the cumulative number of minutes per night and the cumulative percentage of the night spent in the supine posture by 24.7 minutes and 14.7%, respectively.

For the bed partners, compared to video-based posture, the NightOwl overestimated the cumulative percentage of the night spent in the supine posture by 6.4%. The NightOwl also overestimated the cumulative number of minutes per night and the cumulative percentage of the night spent in the prone posture by 17.3 minutes and 7.4%, respectively.

These discrepancies between the video-based posture and NightOwl-based posture are mostly explained by the differing classification systems they use (previously explained in the Discussion section). Briefly, the NightOwl uses a five-posture classification system, whereas the video analysis uses a thirteen-posture classification system. We did not have access to the NightOwl's raw accelerometry data nor were we privy to the angles of lateral tilt at which the NightOwl classified a posture as left, supine, right, or prone. For example, when a participant was in the left tilt posture as determined by video analysis, the NightOwl may classify them in the left lateral posture or the supine posture, depending on the angle of lateral tilt it detects (we observed this in one participant, for example, whose right hip, pelvis, back, and shoulder was on top of her pregnant pillow and was, therefore, clearly in a left tilt posture; however, the NightOwl classified her posture as supine). Given this, comparison of measurements from a five-posture to a thirteen-posture classification system is difficult (and an apples-to-oranges comparison per se). As an example, consider the pregnant participants: if the mean (not medians as shown in **Table S6**) cumulative number of minutes and cumulative percentage of the night spent in the supine and six *near-supine* postures (i.e., left tilt, supine thorax with left pelvic tilt, supine thorax with right pelvic tilt, supine pelvis with left thorax tilt, supine pelvis with right thorax tilt, and right tilt) are added up, the sum is 96.2 minutes and 23.7%, respectively, which is significantly more aligned with the NightOwl's "supine" determination (mean 70.2 minutes and mean 24.6%, respectively; data not shown).

Other reasons, albeit less significant, for discrepancies between the video-based posture and NightOwl-based posture are with regard to placement of the NightOwl position sensor just below the xiphoid process on the person's abdomen. While scoring the video, on several occasions, we noticed that the NightOwl was donned upside down, which reversed the polarity of the NightOwl's "left" and "right" classifications. Furthermore, we also noticed that the NightOwl was donned sideways on one participant one night, which resulted in the NightOwl classifying "prone" and "supine" postures as "left" and "right", and vice versa. For another person in which we noticed that the NightOwl posture classifications were awry for one night, careful review of the video while scoring it showed (during a moment around 3:00 AM when the person change postures and the bed sheets temporarily were not obscuring the body) that the NightOwl position sensor, while initially donned correctly on the abdomen, had become detached from the correct location and reattached on the left side of the back. We also observed at least one couple who somehow donned each other's NightOwl position sensors for one night, which resulted in the NightOwl's posture classifications for the pregnant participant actually being the posture classifications for the bed partner, and vice versa. Finally, particular to the discrepancy between the video-based posture and NightOwl-based posture in the cumulative percentage of the night spent in each posture, the denominators of the percentage calculation are different, which the video-based calculation using the total time in bed but the NightOwl-based calculation using the total sleep time.

Table S6 | **Cumulative number of minutes per night and cumulative percentage of the night spent in each posture for the pregnant participant and bed partner based on video analysis and NightOwl data**

| | **Pregnant** (n=38) | **Bed Partner** (n=31) |
|---|---|---|

| | Video based | NightOwl based | **Video - NO** Difference 95% CI p-value | Video based | NightOwl based | **Video - NO** Difference 95% CI p-value |
|---|---|---|---|---|---|---|
| *Left recovery* | | | | | | |
| Minutes | 2.4 (18.0) | n/a | n/a | 21.1 (56.9) | n/a | n/a |
| Percent of night[a] | 0.5 (4.5) | n/a | n/a | 8.4 (14.8) | n/a | n/a |
| *Left lateral* | | | | | | |
| Minutes | 148.0 ± 61.0 | 125.5 ± 70.2 [3] | 18.9 (0.26 to 37.5) **0.047** | 69.0 (67.2) | 83.4 ± 56.6 [8] | 5.8 (-27.8 to 27.0) 0.60 |
| Percent of night[a] | 38.0 ± 17.6 | 39.2 ± 20.4 [3] | -1.4 (-7.8 to 4.9) 0.65 | 16.4 (19.4) | 26.7 ± 15.6 [8] | -2.2 (-9.7 to 2.8) 0.27 |
| *Left tilt* | | | | | | |
| Minutes | 11.5 (40.8) | n/a | n/a | 1.6 (5.1) | n/a | n/a |
| Percent of night[a] | 2.6 (9.8) | n/a | n/a | 0.42 (1.1) | n/a | n/a |
| *Supine* | | | | | | |
| Minutes | 17.7 (47.0) | 52.0 (86.1) [3] | -24.7 (-39.9 to -11.1) **<0.001** | 81.9 (117.4) | 69.4 (138.7) [8] | 2.9 (-17.4 to 19.0) 0.69 |
| Percent of night[a] | 3.8 (12.9) | 20.0 (35.3) [3] | -14.7 (-19.3 to -7.9) **<0.001** | 20.7 (23.6) | 21.2 (42.7) [8] | -6.4 (-11.8 to -1.9) **0.007** |
| *STRP* | | | | | | |
| Minutes | 1.0 (7.1) | n/a | n/a | 0.10 (2.4) | n/a | n/a |
| Percent of night[a] | 0.2 (1.7) | n/a | n/a | 0.02 (0.86) | n/a | n/a |
| *STLP* | | | | | | |
| Minutes | 0.2 (6.0) | n/a | n/a | 0.43 (4.1) | n/a | n/a |
| Percent of night[a] | 0.05 (1.3) | n/a | n/a | 0.10 (1.0) | n/a | n/a |
| *SPRT* | | | | | | |
| Minutes | 0 (0) | n/a | n/a | 0 (0) | n/a | n/a |
| Percent of night[a] | 0 (0) | n/a | n/a | 0 (0) | n/a | n/a |
| *SPLT* | | | | | | |
| Minutes | 0 (0) | n/a | n/a | 0 (0) | n/a | n/a |
| Percent of night[a] | 0 (0) | n/a | n/a | 0 (0) | n/a | n/a |
| *Right tilt* | | | | | | |
| Minutes | 5.6 (22.6) | n/a | n/a | 2.7 (15.8) | n/a | n/a |
| Percent of night[a] | 1.2 (4.8) | n/a | n/a | 0.66 (3.2) | n/a | n/a |
| *Right lateral* | | | | | | |
| Minutes | 125.8 ± 80.6 | 96.1 ± 65.6 [3] | 27.1 (4.6 to 49.5) **0.02** | 77.3 (58.1) | 77.1 ± 42.9 [8] | 5.7 (-17.1 to 31.7) 0.64 |
| Percent of night[a] | 29.4 ± 14.8 | 31.9 ± 20.2 [3] | -3.0 (-8.4 to 2.3) 0.26 | 19.6 (16.5) | 24.5 ± 14.9 [8] | -2.7 (-10.1 to 4.7) 0.43 |
| *Right recovery* | | | | | | |
| Minutes | 2.1 (19.4) | n/a | n/a | 30.1 (70.5) | n/a | n/a |
| Percent of night[a] | 0.5 (4.7) | n/a | n/a | 6.4 (14.9) | n/a | n/a |
| *Prone* | | | | | | |
| Minutes | 0 (0) | 0.0 (3.3) [3] | n/a | 12.5 (36.4) | 35.0 (62.3) | -17.3 |

| | | | | | | |
|---|---|---|---|---|---|---|
| | | | | | [8] | (-32.3 to -7.1) **<0.001** |
| Percent of night[a] | 0 (0) | 0 (1.5) [3] | n/a | 2.6 (11.7) | 9.8 (22.5) [8] | -7.4 (-20.4 to -2.9) **<0.001** |
| *Sitting* | | | | | | |
| Minutes | 0.3 (1.4) | n/a | n/a | 0.17 (0.71) | n/a | n/a |
| Percent of night[a] | 0.08 (0.4) | n/a | n/a | 0.07 (0.18) | n/a | n/a |

*p<0.05, **p<0.01, ***p<0.001
†The 95% CI is given if the difference is statistically significant.
[a]For video-based values, the denominator for the percentage calculation is the length of time in bed, whereas for the Night-Owl-based values the denominator is the total sleep time.
Normally distributed variables are presented as mean ± standard deviation and difference testing is by paired samples Student T-test when both samples are normally distributed. Non-normally distributed variables are presented as median (interquartile range) and difference testing is by paired samples Wilcoxon signed-rank test if one or both samples have a non-normal distribution. Square brackets with a number enclosed, [number], indicates the number of missing values.
**Abbreviations**: NO indicates NightOwl; CI indicates confidence interval. STRP indicates supine thorax, right pelvis; STLP indicates supine thorax, left pelvis; SPRT indicates supine pelvis, right thorax; SPLT indicates supine pelvis, left thorax.

#### Sleeping Posture Changes: With and Without Bed Partner

Overnight videos were analyzed to determine, for the pregnant participants with bed partners (n=33) and the pregnant participants without bed partners (n=5; note that all five of these participants normally slept with their bed partner, but their bed partner opted out from participating in the study), the average time in bed per night, the total number of posture changes per person per night, the total nightly posture change index (PCI; the number of posture changes occurring per hour), and the hour-by-hour PCI from the first hour through the last hour of the overnight video recordings to show how the frequency of posture changes varies throughout the night (see **Table S7**). Participants (and their bed partner, if applicable) who completed the sleep studies postpartum were excluded from this analysis. Pregnant participants who slept with their bed partner changed posture less times per night than those who slept without their bed partner, but this did not reach SS. Except for hour 7, there was a trend of the PCI of pregnant participants sleeping with their bed partner being less than those sleeping without their bed partner, but this did not reach SS. These results (less posture changes when sleeping with one's bed partner, and more posture changes when sleeping without one's bed partner) indicate less stable and more restless sleep when sleeping without one's bed partner and are aligned with evidence that partner presence functions as a safety signal, which reduces stress and promotes better sleep.[1–4] For example, Drews et al. showed improved sleep (more REM sleep, less fragmented REM sleep, longer undisturbed REM fragments) when couples slept together compared to when they slept separately.[5] A limitation of this analysis, however, is the low sample size in the group of pregnant participants who slept without their bed partner, which meant we were underpowered to detect a smaller differences.

Table S7 | **Average time in bed per night, number of posture changes per person per night, total posture change index, and hour-by-hour posture change index for the pregnant participants sleeping with and without their bed partner based on video analysis**

| | **Pregnant participants sleeping with their bed partner** (n=33) | **Pregnant participants sleeping without their bed partner** (n=5) | **With bed partner – Without bed partner** Difference (95% CI) | p-value |
|---|---|---|---|---|
| Average time in bed per night (hours) | 7.0 ± 1.7 | 6.9 ± 0.68 | 0.18 (-1.4 to 1.8) | 0.82 |
| Total number of posture changes per person per night | 23.3 ± 10.3 | 28.4 ± 10.9 | -5.1 (-15.1 to 5.0) | 0.31 |
| *Posture change index* | | | | |
| Total | 3.0 (1.6) | 4.3 ± 1.6 | -1.3 (-2.5 to 0.80) | 0.24 |
| First hour | 4.0 (2.8) | 5.2 ± 3.8 | -0.42 (-5.0 to 2.5) | 0.83 |
| Hour 2 | 2.0 (2.0) | 2.9 ± 2.2 | -0.33 (-2.9 to 1.5) | 0.78 |
| Hour 3 | 3.3 ± 1.8 | 4.8 ± 1.7 | -1.5 (-3.2 to 0.27) | 0.09 |
| Hour 4 | 3.4 (3.0) | 3.7 ± 2.3 | -0.6 (-2.5 to 1.8) | 0.56 |
| Hour 5 | 3.4 ± 1.9 [1] | 4.5 ± 1.0 | -1.1 (-2.9 to 0.67) | 0.22 |
| Hour 6 | 2.9 (1.8) [1] | 4.2 ± 0.93 | -1.1 (-2.3 to 0.50) | 0.10 |
| Hour 7 | 3.2 (2.8) [3] | 2.7 ± 1.8 | 0.72 (-1.3 to 3.5) | 0.51 |
| Hour 8 | 3.0 ± 1.6 [7] | 4.0 ± 2.7 | -1.0 (-2.8 to 0.82) | 0.27 |
| Hour 9 | 3.0 (3.2) [10] | 7.1 ± 1.2 [1] | -3.7 (-5.4 to 2.0) | 0.09 |
| Hour 10 | 3.3 ± 2.1 [24] | n/a [5] | n/a | n/a |
| Hour 11 | 5.3 (1.9) | n/a | n/a | n/a |

| | | |
|---|---|---|
| | [27] | [5] |

Normally distributed variables are presented as mean ± standard deviation and difference testing is by Student T-test when both samples are normally distributed. Non-normally distributed variables are presented as median (interquartile range) and difference testing is by Mann-Whitney test if one or both samples have a non-normal distribution. Square brackets with a number enclosed, [number], indicates the number of missing values.
**Abbreviations**: CI indicates confidence interval.

### References


1. Elsey T, Keller PS, El-Sheikh M. The role of couple sleep concordance in sleep quality: Attachment as a moderator of associations. *J Sleep Res*. 2019;28(5):e12825. doi:10.1111/jsr.12825

2. Doerr JM, Klaus K, Troxel W, et al. The Effect of Intranasal Oxytocin on the Association Between Couple Interaction and Sleep: A Placebo-Controlled Study. *Psychosom Med*. 2022;84(6):727-737. doi:10.1097/PSY.0000000000001091

3. Troxel WM, Buysse DJ, Matthews KA, et al. Marital/cohabitation status and history in relation to sleep in midlife women. *Sleep*. 2010;33(7):973-981. doi:10.1093/sleep/33.7.973

4. Drews HJ, Drews A. Couple Relationships Are Associated With Increased REM Sleep-A Proof-of-Concept Analysis of a Large Dataset Using Ambulatory Polysomnography. *Front Psychiatry*. 2021;12:641102. doi:10.3389/fpsyt.2021.641102

5. Drews HJ, Wallot S, Brysch P, et al. Bed-Sharing in Couples Is Associated With Increased and Stabilized REM Sleep and Sleep-Stage Synchronization. *Front Psychiatry*. 2020;11:583. doi:10.3389/fpsyt.2020.00583
